# Analysis of Covid-19 Data for Eight European Countries and the United Kingdom Using a Simplified SIR Model

**DOI:** 10.1101/2020.05.26.20114058

**Authors:** Gyan Bhanot, Charles DeLisi

**Affiliations:** Department of Physics and Astronomy, Rutgers University, Piscataway, NJ, 08854, USA; Department of Molecular Biology and Biochemistry, Rutgers University, Piscataway, NJ, 08854, USA; Rutgers Cancer Institute of New Jersey, New Brunswick, NJ, 08903, USA; Moores Cancer Center, University of California San Diego, La Jolla, CA 92103, USA; Department of Biomedical Engineering, Boston University, Boston, MA 02215, USA

**Keywords:** Epidemic modelling, SARS-Cov-2/Covid-19 pandemic, SIR model, predictions

## Abstract

Understanding the characteristics of the SARS-Cov-2/Covid-19 pandemic is central to developing control strategies. Here we show how a simple Susceptible-Infective-Recovered (SIR) model applied to data for eight European countries and the United Kingdom (UK) can be used to forecast the descending limb (post-peak) of confirmed cases and deaths as a function of time, and predict the duration of the pandemic once it has peaked, by estimating and fixing parameters using only characteristics of the ascending limb and the magnitude of the first peak. As with all epidemiological analyses, unanticipated behavioral changes will result in deviations between projection and observation. This is abundantly clear for the current pandemic. Nonetheless, accurate short-term projections are possible, and the methodology we present is a useful addition to the epidemiologist’s armamentarium. Since our predictions assume that control measures such as lockdown, social distancing, use of masks etc. remain the same post-peak as before peak, deviations from our predictions are a measure of the extent to which loosening of control measures have impacted case-loads and deaths since the first peak and initial decline in daily cases and deaths. The predicted and actual case fatality ratio, or number of deaths per million population from the start of the pandemic to when daily deaths number less than five for the first time, was lowest in Norway (pred: 44 ± 5 deaths/million; actual: 36 deaths/million) and highest for the United Kingdom (pred: 578 +/- 65 deaths/million; actual 621 deaths/million). The inferred pandemic characteristics separated into two distinct groups: those that are largely invariant across countries, and those that are highly variable. Among the former is the infective period, 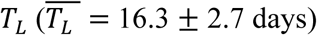; the average time between contacts, 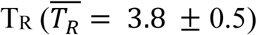 and the average number of contacts while infective, 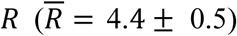. In contrast, there is a highly variable time lag *T*_*D*_ between the peak in the daily number of confirmed cases and the peak in the daily number of deaths, ranging from a low of *T*_*D*_ = 2,4 days for Denmark and Italy respectively, to highs of *T*_*D*_ = 12, 15 for Germany and Norway respectively. The mortality fraction, or ratio of deaths to confirmed cases, was also highly variable, ranging from low values 3%, 5% and 5% for Norway, Denmark and Germany respectively, to high values of 18%, 20% and 21% for Sweden, France, and the UK respectively. The probability of mortality rather than recovery was a significant correlate of the duration of the pandemic, defined as the time from 12/31/2019 to when the number of daily deaths fell below 5. Finally, we observed a small but detectable effect of average temperature on the probability *α* of infection per contact, with higher temperatures associated with lower infectivity. Policy implications of our findings are also briefly discussed.

## I. Introduction

Coronaviruses are large, enveloped, single-stranded RNA viruses which are widespread in animals and usually cause only mild respiratory illnesses in humans [1-5]. However, in 2003, a new coronavirus SARS-CoV emerged, which caused a life-threatening respiratory disease, with a fatality rate of almost 10% [6,7]. Unfortunately, after an initial burst of interest in development of treatment options, interest in this virus waned. The emergence of a novel coronavirus SARS-CoV-2, identified in January 2020 as the likely causative agent of a cluster of pneumonia cases which first appeared in Wuhan, China in December 2019, has since caused a worldwide pandemic [8-13]. SARS-CoV-2 is the seventh known coronavirus to cause pathology in humans [1]. The associated respiratory illness, called COVID-19, ranges in severity from a symptomless infection [8], to common-cold like symptoms, to viral pneumonia, organ failure, neurological complications and death [9-11]. While the mortality in SARS-CoV-2 infections is lower than in SARS-CoV [9-12], it has more favorable transmission characteristics, a higher reproduction number [13], and as we will show, a long latency period and asymptomatic infective phase.

The governments of several countries have taken significant measures to slow the infection rate of Covid-19, such as social distancing, quarantine, identification, tracking and isolation. However, there is no uniform policy, some governments have reacted later than others and some (e.g. Sweden) made a deliberate decision to keep the country open, leaving counter-measures up to individuals.

A large amount of consistent public data is now available on the number of tests performed, the number of confirmed infected cases, and the number of deaths in different contexts, such as locations and health conditions [14]. These provide important sources of information for the development and testing of models that can identify pandemic characteristics affecting viral dynamics, and guide public policy by predicting the impact of various interventions [15].

All data, of course, have limitations, and it’s ultimately the completeness and quality of data that limit the success of models. It is well known that confirmed infected cases seriously underestimate the actual number of infections [16,17]: not everyone who is infected is symptomatic, and not everyone who dies from the disease has been tested [18]. Even the number of reported deaths may be underestimated because of co-mortalities; i.e. COVID-19 increases susceptibility to other diseases and conditions [19]. Moreover, the virus can be transmitted by asymptomatic individuals – who comprise a substantial portion of the infected population [20] —militating against accurate estimates of transmission probability. Nonetheless as indicated in [21] and by our own verified forecasts, models can provide useful information.

Dynamical (mechanistic) models, such as the one presented here, have been used for forecasting (meaning that once initial conditions are set, there are no changes in the model) and for projecting (the outcome is changed by intervention strategies). For example, projections and forecasting models of various types have been used as early as February, 2020 to determine a reproductive number [13]. More generally multiple research groups have used them to estimate Case Fatality Ratios (CFRs) [22], to forecast and project the need for hospital beds [23] and to project and forecast mortality [24]. More specifically, among the many applications to COVID-19, four variable Susceptible-Exposed-Infective-Recovered models have been used to project the impact of social distancing on mortality [25], three variable Susceptible-Infective-Recovered models to estimate case fatality and recovery ratios early in the pandemic [26], and a time delayed SIR has been used to evaluate the effectiveness of suppression strategies [27]. One of the most ambitious dynamical models, which includes 8 state variables and 16 parameters was fruitfully applied to evaluate intervention strategies in Italy, in spite of the fact that parameter identifiability could not be assured [28].

Our simplified version of the original SIR model [29] differentiates itself from the studies done so far by a unique methodological approach to rigorous identification of parameters, and by making a number of useful predictions very easily, such as the duration of the pandemic in different countries and case fatality ratios, using only characteristics of the increasing portion of verified cases vs time, and the amplitude of the peak in the daily cases. Although it is sometimes said that using daily mortality data is more reliable than daily verified cases, we find that at least in the countries we analyzed, the patterns for these two quantities are virtually identical, differing only in amplitude and a rigid time translation.

It is important to note that extrapolating our model predictions for daily cases and deaths past the peak assumes that control measures such as lockdown, social distancing, use of masks etc and care given to Covid-19 patients remained the same after the peak in daily cases as before the peak. Observed deviations from our predictions are a measure of the extent to which loosening of control measures or changes in care of infected individuals impacted case-loads and deaths past the peaks in the number of cases and deaths.

## II. Methods

We model the Covid-19 pandemic using a simplified version of the SIR model [29], which partitions the population into three compartments, Susceptibles (S), Infectious (I) and Removed R: Recovered or Dead after being infected. This and other models to study the global spread of diseases have been used in a variety of contexts (For some recent reviews, see [30-32]).

So far, the Covid-19 pandemic, at least in the developed countries in Europe where we will apply this model, seems to have the following dynamics: After being infected, an individual remains able to infect others for an average of *T*_*L*_ days. After a time *T*_*L*_, the infected individual becomes sick, gets tested, is identified as infected and is removed from the pool by quarantine or hospitalization. Thus, in our context, the SIR model dynamics can be defined as follows: At t=0, there is a pool of *N* interacting individuals, almost all of whom are in the S compartment, except for the few infected cases in the I compartment. The R compartment is empty at t=0. Over time, individuals move from S to I and from I to R. In R, they either recover or die. Since the Recovered pool is populated only from the Infected pool, on average, the number removed each day must equal the number infected sometime in the past; i.e. the two are related by a fixed time displacement and a “mortality probability” factor. We assume that the number of deaths and the number of individuals recovered each day are proportional to the number Removed each day by fixed probabilities, that remain invariant over the course of the epidemic; i.e. that the number dead or recovered each day are proportional to the number infected on some previous day, with different time delays and probabilities.

### The Model

Let,

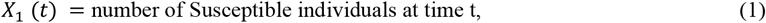

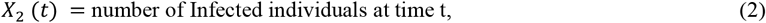

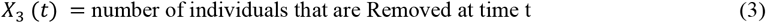

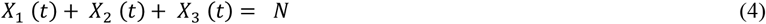

A fraction *δ* of the infected individuals will die after being identified as infected. On average, there will be a time delay *T*_*D*_ between when a person is identified to be infected (tests positive) and when he/she dies of the disease. *T*_*D*_ will depend on a variety of factors, such as quality of care, age, severity of disease, co-morbidities, immune status etc.

Under these assumptions, the number of deaths *X*_4_(*t*) at time *t* is related to the number of Infected cases *X*_2_ by:

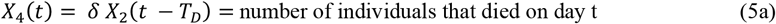

Similarly, the number of Recovered at time *t* will be:

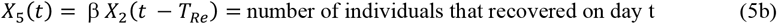

Let,

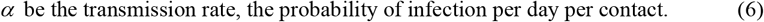

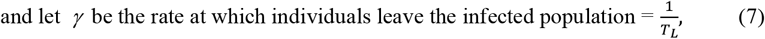

with *T*_*L*_ being interpreted as the latency, or the average time interval during which an infected individual can infect a naive individual.

The equations governing the dynamics are:

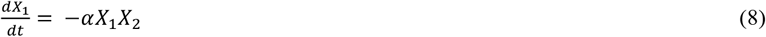

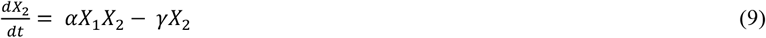

The initial conditions at *t* = 0 are:

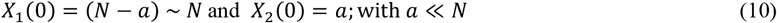

An equation relating the state variables *X*_1_ and *X*_2_ can be obtained by dividing (8) by (9) and integrating. This gives,

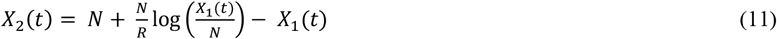

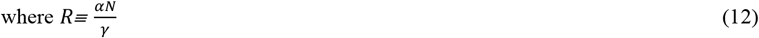

Hence,

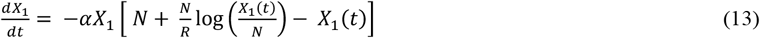

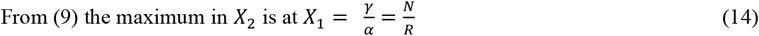

Substituting this into (13) gives:

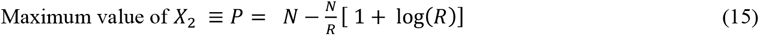

At *t* = ∞, *X*_2_ = 0.Hence, from (13), we get:

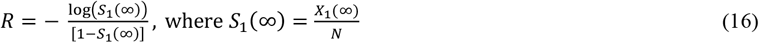

as the fraction of susceptible individuals at t = ∞.

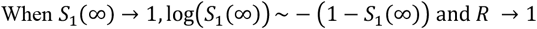

This says that when nobody is infected, *S*_1_(∞) = 1, i.e. there is no pandemic. This corresponds to *R* = 1. It is easy to show that if *R* < 1, there are no solutions to (16) that satisfy 0 ≤ *S*_1_(∞) ≤ 1.

### Fitting the Model to Data

We focus on four North European countries, Netherlands, Denmark, Sweden and Norway, denoted by EN, four South European countries, France, Italy, Spain and Germany, denoted by ES, and the United Kingdom (UK). Data for the number of confirmed cases and deaths were obtained from https://ourworldindata.org/coronavirus-source-data, from an EU agency established in 2005 and based in Stockholm with the aim to strengthen Europe’s defense against infectious diseases. The data for the number of tests was obtained from https://ourworldindata.org/coronavirus-testing [33].

We note that the data identify the daily number of Confirmed cases, whereas the SIR model we described above requires the number of Infected cases. However, the number of confirmed cases at time t are derived from the infected cases at some previous time t’, where the time lag between t and t’ is fixed but unknown. Since Equations (1-5) are invariant under a time translation, we can use the data for daily confirmed cases to represent *X*_2_ with the understanding that there is an implicit shift in time between a person becoming infected and being identified as such in the data. This shift in time would depend on how quickly infected individuals are identified and included in the case count, and hence may vary from country to country. However, we assume that within a given country, this time interval is fixed on average. With this caveat, in the results below, we will use the confirmed daily cases in our analysis to represent *X*_2_(*t*), and use either the term “infected” or “confirmed” when discussing *X*_2_.

The first date for which data was available for these countries was 12/31/2019, which we denote as day number 0 in our analysis and in the plots to follow. Among the countries considered here, the earliest cases were identified in France on 1/25/2020, which corresponds to day 25. In the other countries, the earliest cases were identified on the following days, counting from 12/31/2019: Netherlands: day 59, Denmark: day 58, Sweden: day 32, Norway: day 58, UK: day 31, Spain: day 32, Germany: day 28, Italy: day 31.

The available data includes

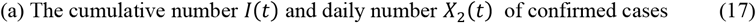

and

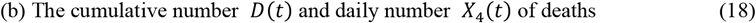

These are related by:

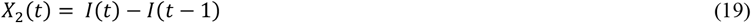

and

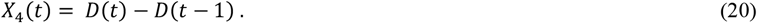

It is worth noting that the dynamics of the number removed each day and the total number of removed do not enter our analysis explicitly. Some connection to the full SIR model [29] can be made by noting that the total number of removed individuals increases asymptotically at large times to *N*(1 − *S*_1_(∞)).

We determine and analyze the following parameters country by country: ***N***, as defined in (4); ***α***, the transmission rate; i.e. the number of infections per day per contact; ***γ***, the average rate at which individuals leave the infected pool; ***R***, the average number of transmissions per individual**; *T***_***R***_, the time between transmissions; ***δ***, the fraction of individuals in the infected pool who will, on average, die ***T***_***D***_ days later; β **= 1 – *δ***, the fraction of individuals in the infected pool who will, on average, recover ***T***_***Re***_ days later; and ***T***_***L***_, the infective period. We also determine the duration of the infection, defined as the number of days from 12/31/19 until the number of deaths drops below 5 per day.

The parameters *N, α*, ***γ***, *R* were obtained using (5), (8) and (9) to do numerical fits. Because of the definition of R (see (12)), only three of the parameters, *N, α*, ***γ***, *R*, need to be determined from the data. Consequently, we proceed as follows:

1. Using (12), we define *α* in terms of *N*, ***γ***, *R*. This eliminates *α*.
2. Estimating *P* = maximum value of *X*_2_(*t*) from the data, we determine *N* in terms of *R* using (15). This eliminates *N*.
3. ***γ***(*R* − 1) is determined as the coefficient of *t* in the exponential rise of *X*_2_(*t*) for small t (see Appendix A). This eliminates ***γ***.
4. Using a numerical solver, we vary *R* to fit the observed data for *X*_2_(*t*).

We emphasize that in determining the parameters, we are only using the ascending limb and the peak in the data for *X*_2_(*t*) and use these to predict how *X*_2_(*t*) will evolve in time past the peak. Using the data for *X*_4_(*t*) and the fitted model, we can also determine *T*_*D*_ and *δ* using (5a) by a simple translation and scaling of the fitted model for *X*_2_(*t*).

Furthermore, once *N, α*, ***γ***, *R* are determined, (16) determines 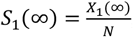, the fraction of the pool of interacting individuals who are naive (uninfected) at the end of the pandemic.

Some other useful parameters we obtain are:

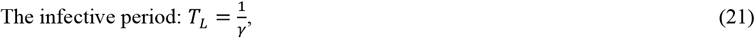

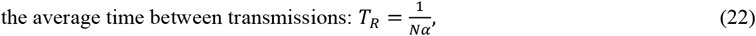

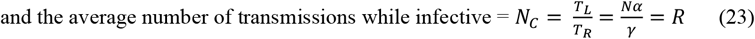

### Data and Fitting Methodology

Although data on deaths is unambiguous, the data for the number of cases is trustworthy only when a sufficient number of tests are performed. It is therefore important to determine whether adequate testing was done to ensure the reliability of case data. Figure 1a and 1b show the cumulative number of tests performed in the countries analyzed, starting from 12/31/2019. We see that in the EN countries, the ratio of cumulative tests to cumulative cases always exceeded five, whereas in the ES countries and the UK, it always exceeded three. Consequently, we expect that the reported number of cases is reliable. It is also important to note that while replicate testing provides some assurance, it leaves unaddressed the problem of quality (reliability), breadth (the percentage of the population reached) and speed (the elapsed time between infection and identification of disease) of testing.

**Figure 1:**
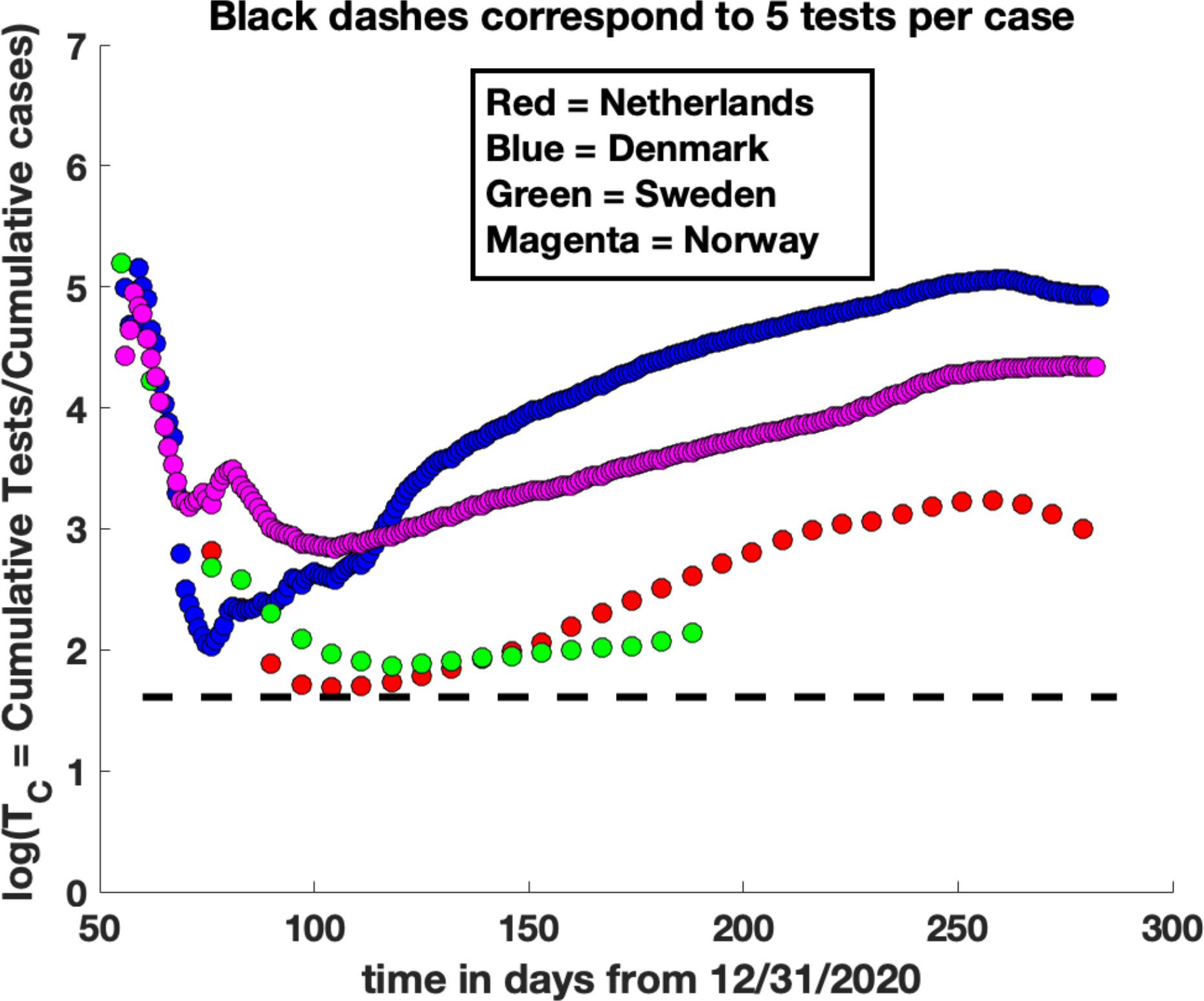
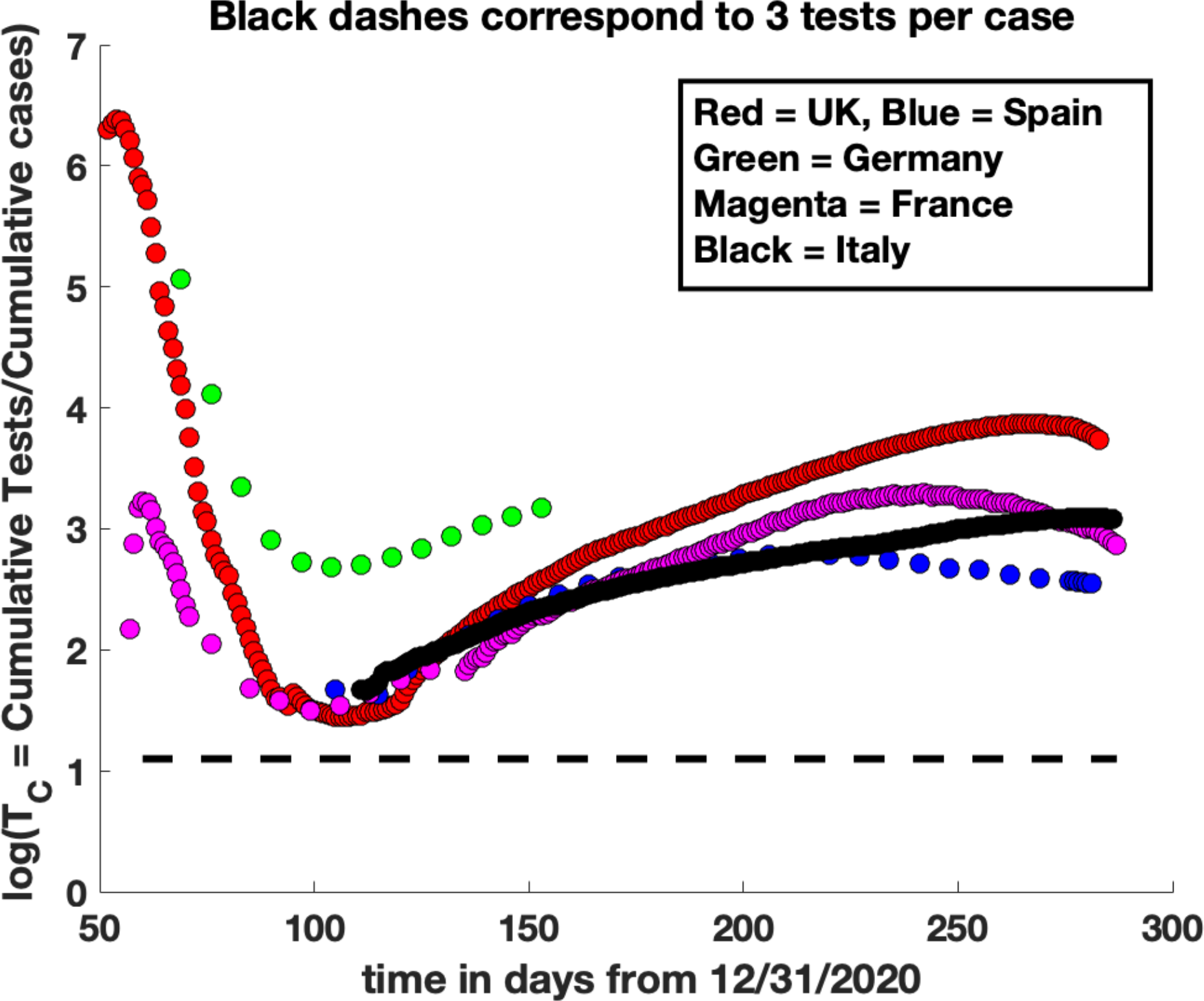
Ratio of the cumulative number of tests to number of identified Covid-19 positive cases as a function of time: **(a)** In the EN countries and **(b)** in the UK and ES countries. The dashed lines in **(a)** and **(b)** represent 5 and 3 tests/case respectively.

One interesting observation from the testing data (Figure 1a, 1b) was that initially, *public concern spread more rapidly than the virus*. As the disease spread and testing ramped up, the initial value of *T*_*C*_ (number of tests/case) was quite large (over 650 in the UK), suggesting that the exponential growth in infections was slower than the exponential public awareness of the disease, resulting in the testing of a large number of people with no disease. Later, as appropriate protocols to identify diseased individuals were established, *T*_*C*_ decreased then flattened out when the testing capacity adequately accounted for new cases, and finally rose again as the pandemic waned.

The values of *X*_2_(*t*) and *X*_4_(*t*) were extracted from the data for the cumulative number of cases and the cumulative number of deaths. To reduce fluctuations in these quantities, the data was averaged over 7 days. Once B = ***γ***(*R* − 1) and P were determined from the exponentially increasing region and the peak in *X*_2_(*t*) respectively (see discussion in previous section), the only undetermined parameter was *R*. Several values of *R* in the range *R* = 1.5 − 6.0 were then tested using the following procedure: For each parameter set, we fitting these data for *X*_2_(*t*) from small t to the peak and beyond, by finding numerical solutions of (8) and (9) using the Matlab Solver *myode2* to determine [*X*_1_(*t*), *X*_2_(*t*)] as a function of time, with the initial conditions, [*X*_1_(*t*_0_) = *N* − *a, X*_2_(*t*_0_) = *a*], starting from a value *t*_0_ of *t* such that *X*_2_(*t*_0_) = *a* ∼ 10.

To determine an error on the fitted parameters, the parameters were varied until a range of values was found that fit the data for *X*_2_(*t*) including fluctuations. Finally, *δ, T_D_* were emperically determined (see eq. (5a)) by scaling and shifting the numerical fits for *X*_2_(*t*) to fit the data for *X*_4_(*t*). Using the fitted parameters, the numerical solutions for [*X*_1_(*t*), *X*_2_(*t*)] were extended to estimate the number of days to the end of the pandemic, *which we define as when the number of daily deaths are less than 5*. The fitted solutions for [*X*_2_(*t*), *X*_4_(*t*)] were also used to compute the predicted total cases per million and the predicted total deaths per million population and compared to their actual values in each country. Finally, the values of *T*_*L*_ and *T*_*R*_ were determined from the fitted data using (21) and (22) respectively.

## III. Results and Discussion

It is important to note that the parameter values we determine are derived solely from: (i) the ascending (small t) limb of *X*_2_(*t*) and (ii) the first peak value of *X*_2_(*t*). Using the fitted parameters to derive results past the peak allows us to: (i) predict how the pandemic would have continued beyond the peak, if control measures such as social distancing, use of PPEs and masks etc. had remained the same as before the peak; (ii) determine deviations of the actual data from our model predictions to estimate the effect of changes in control measures post peak to pandemic dynamics.

The results obtained for *N, α*, ***γ***, *R, δ, T*_*L*_, *T*_*R*_, *T*_*D*_ are given in Tables A. Figures 2 and 3 and Supplementary Figure S1a-i shows the data and fits of our model for *X*_2_ and *X*_4_ respectively. We note that for Sweden and the UK, there is no clear first peak in *X*_2_(*t*) but rather a plateau and a subsequent increase in the number of daily cases. For these countries, we estimated the value of *P* (Eq. 15) from the plateau in Fig. 2c and Fig 2e respectively. For all the other countries, there was a clear first peak in *X*_2_(*t*) whose amplitude was used to estimate P. Unfortunately, for all countries, even those that had a well defined first peak in followed by a decline over time, *X*_2_(*t*) showed significant increase soon after the peak. In some cases, subsequent increases in daily deaths surpassed these first peaks by factors of 2-6 (Figure 1, S1). These suggest that in spite of significant early efforts to control infection rates using social distancing, quarantine, use of masks, testing and other containment measures, there was poor success in limiting the spread of the disease past the first peak in *X*_2_ (Figure 2 a-i), most likely because of a combination of premature lifting of quarantine, inadequate use of facemasks and difficulties in limiting physical contacts, exacerbated by the advent of summer weather and holiday travel.

**Table.**
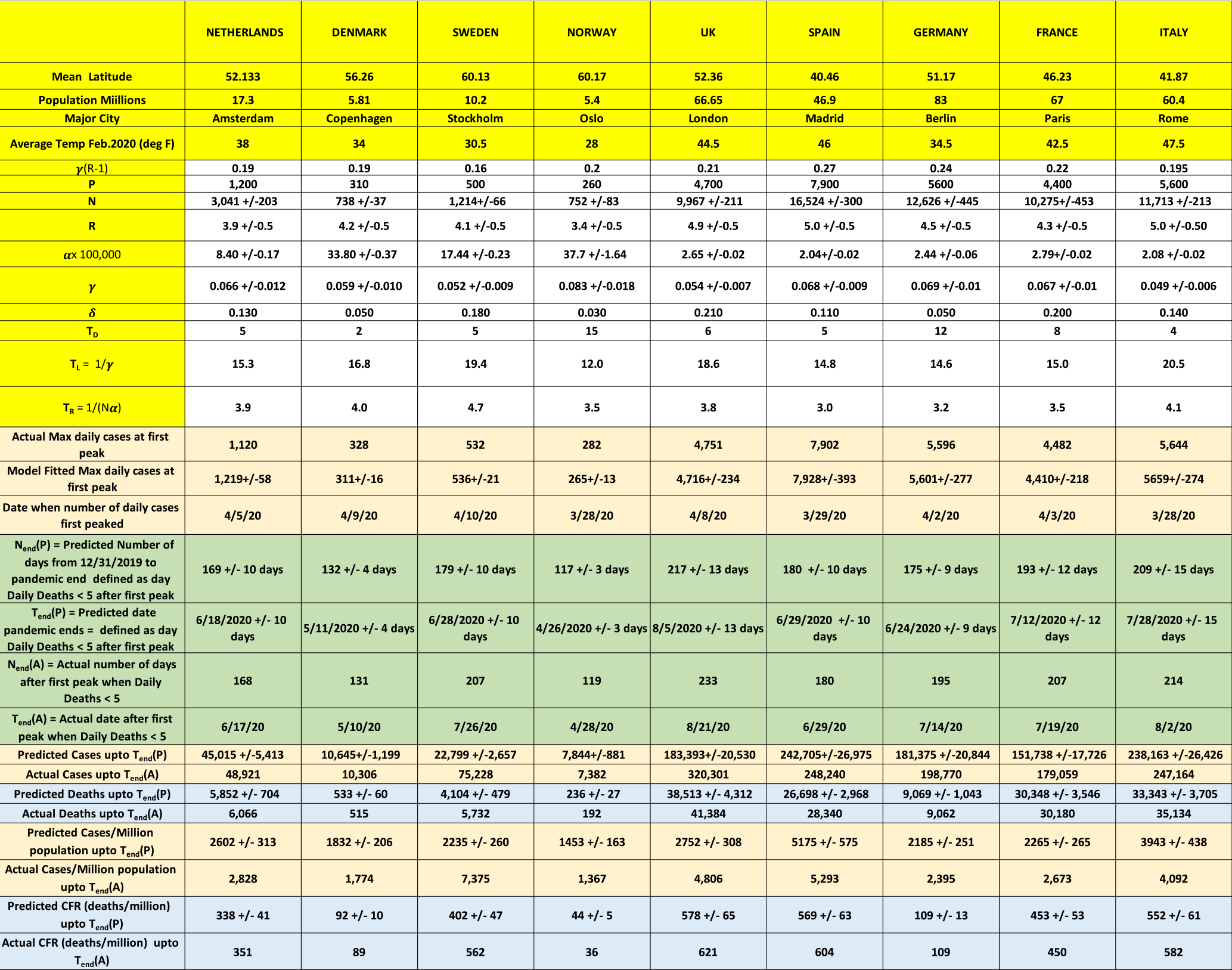
showing a summary of all results for all 9 countries.

**Figure 2:**
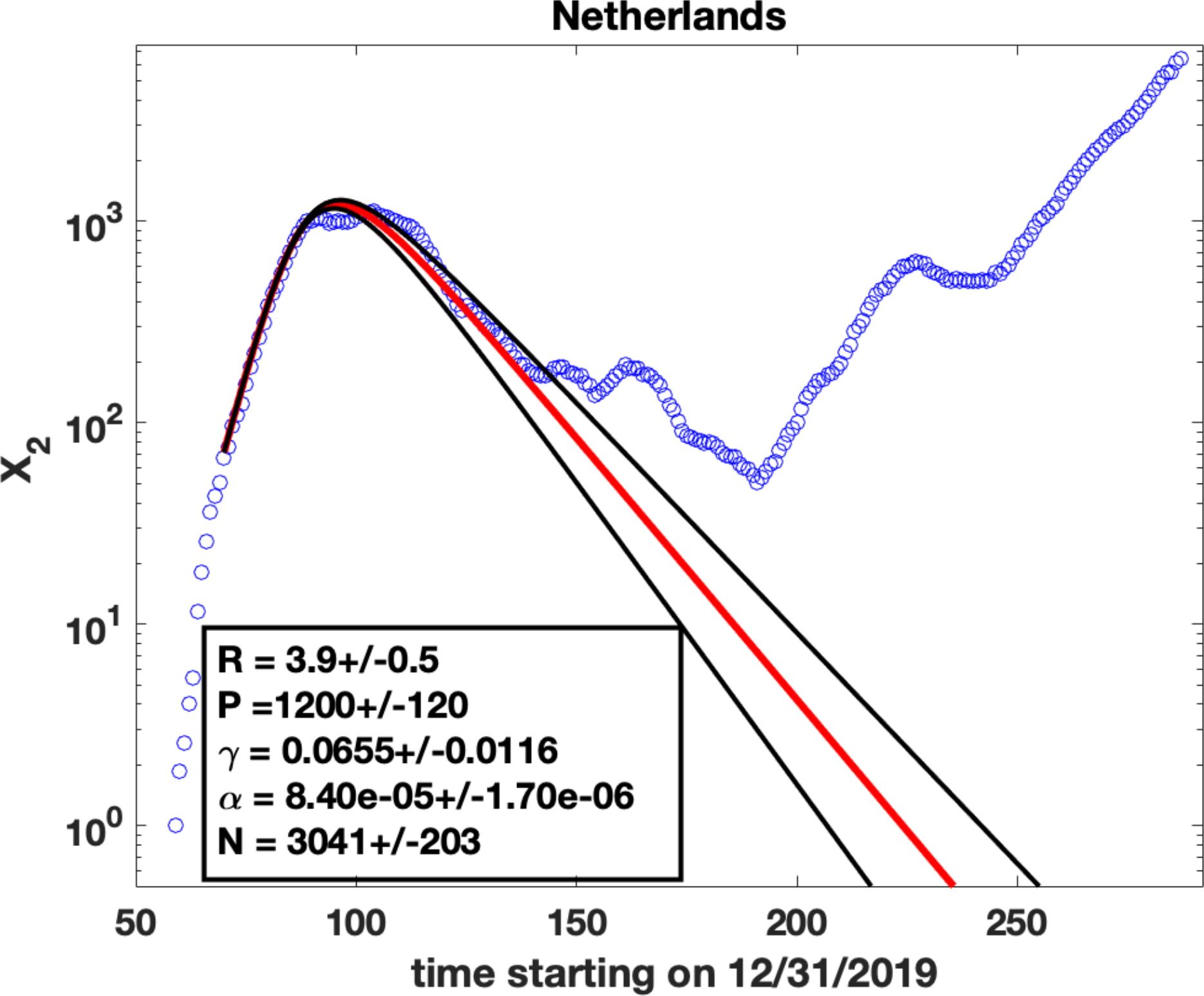
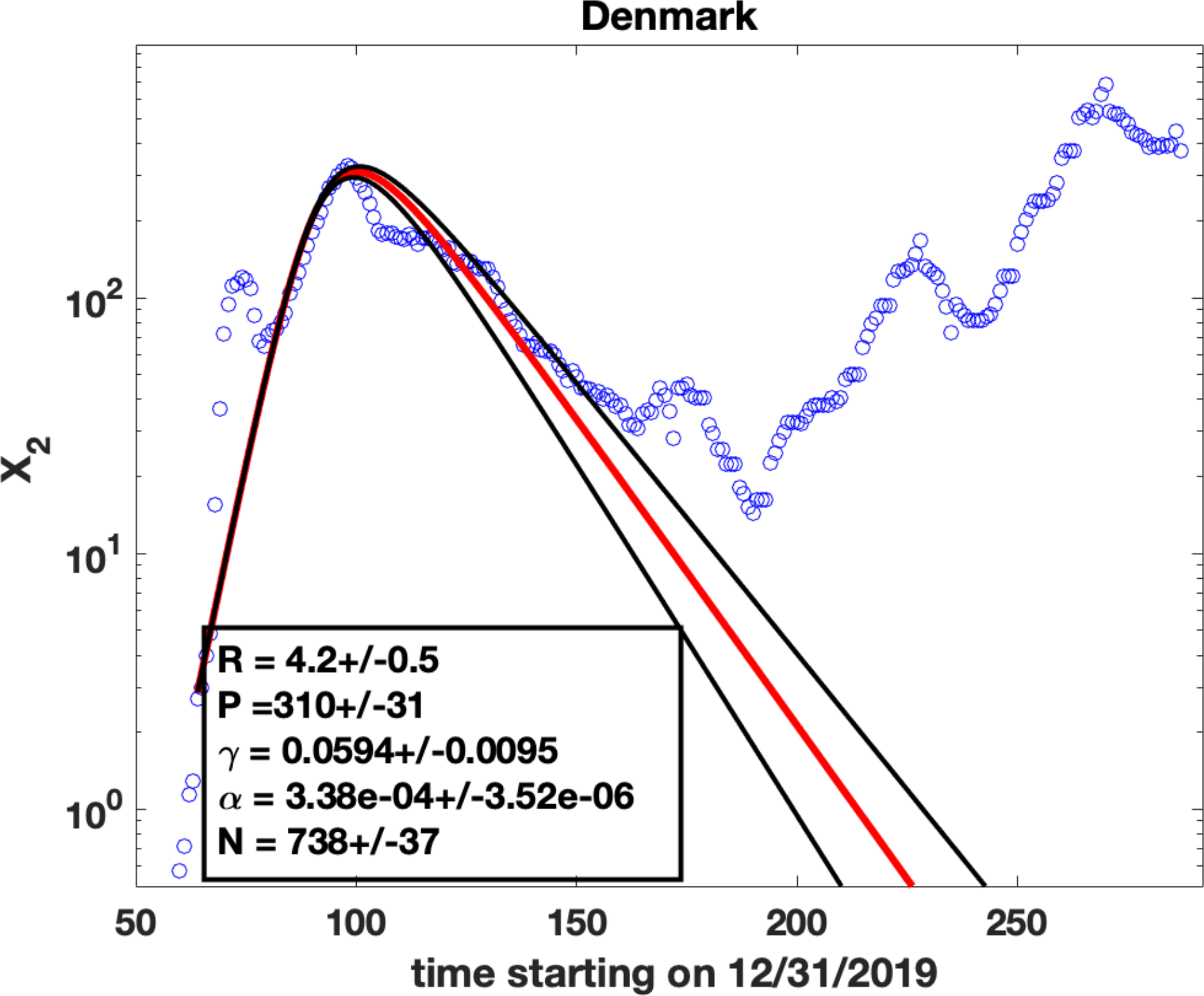
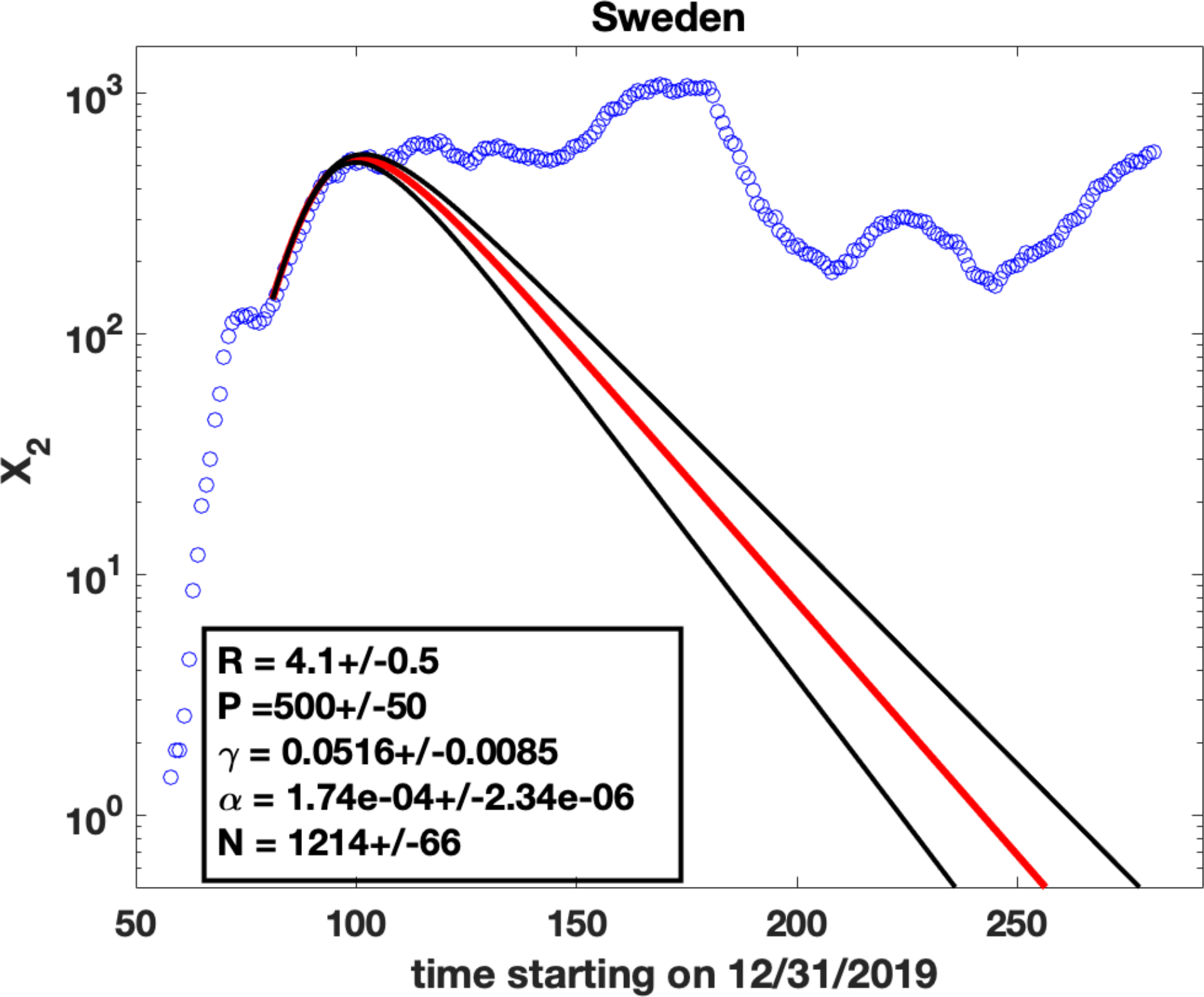
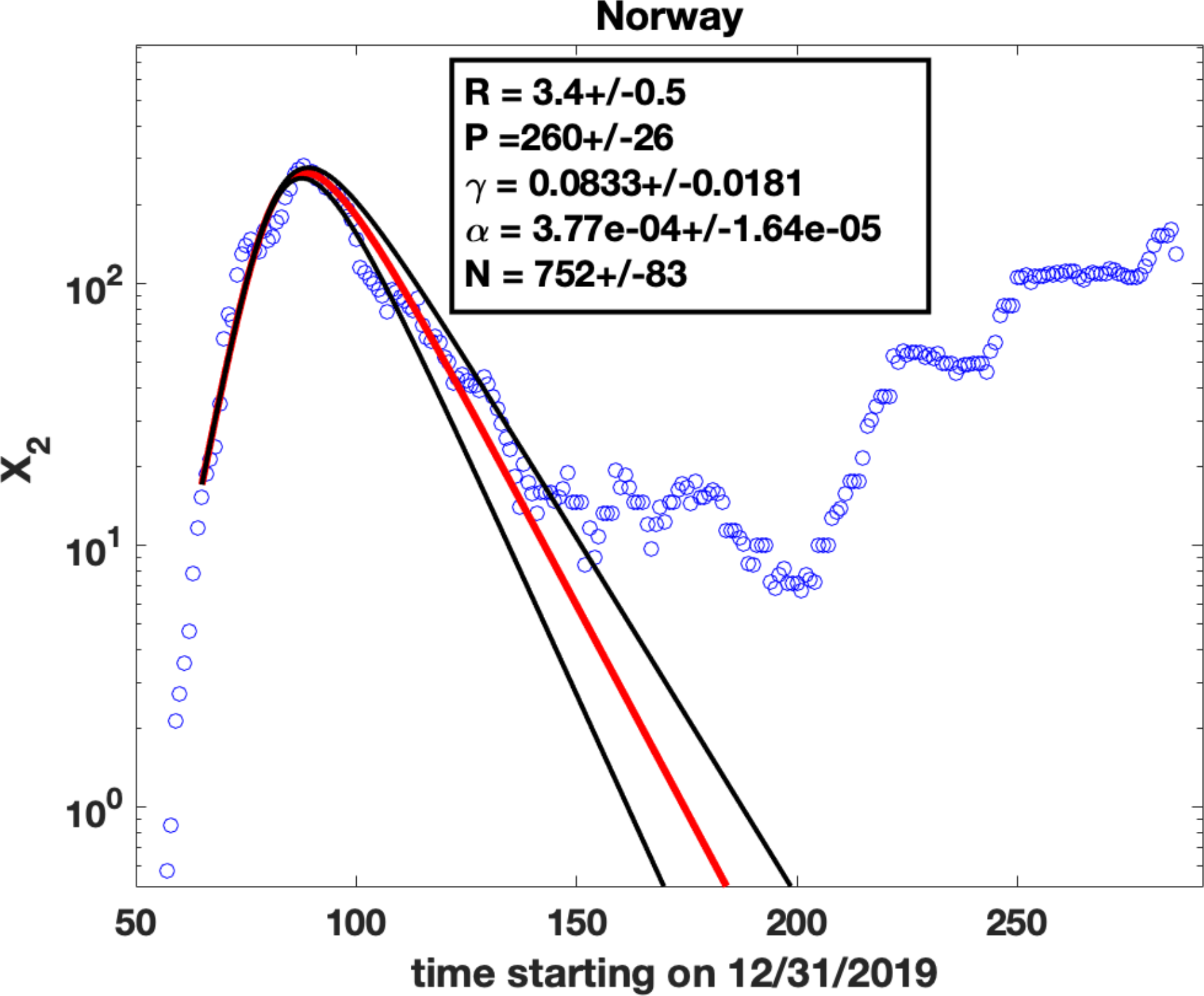
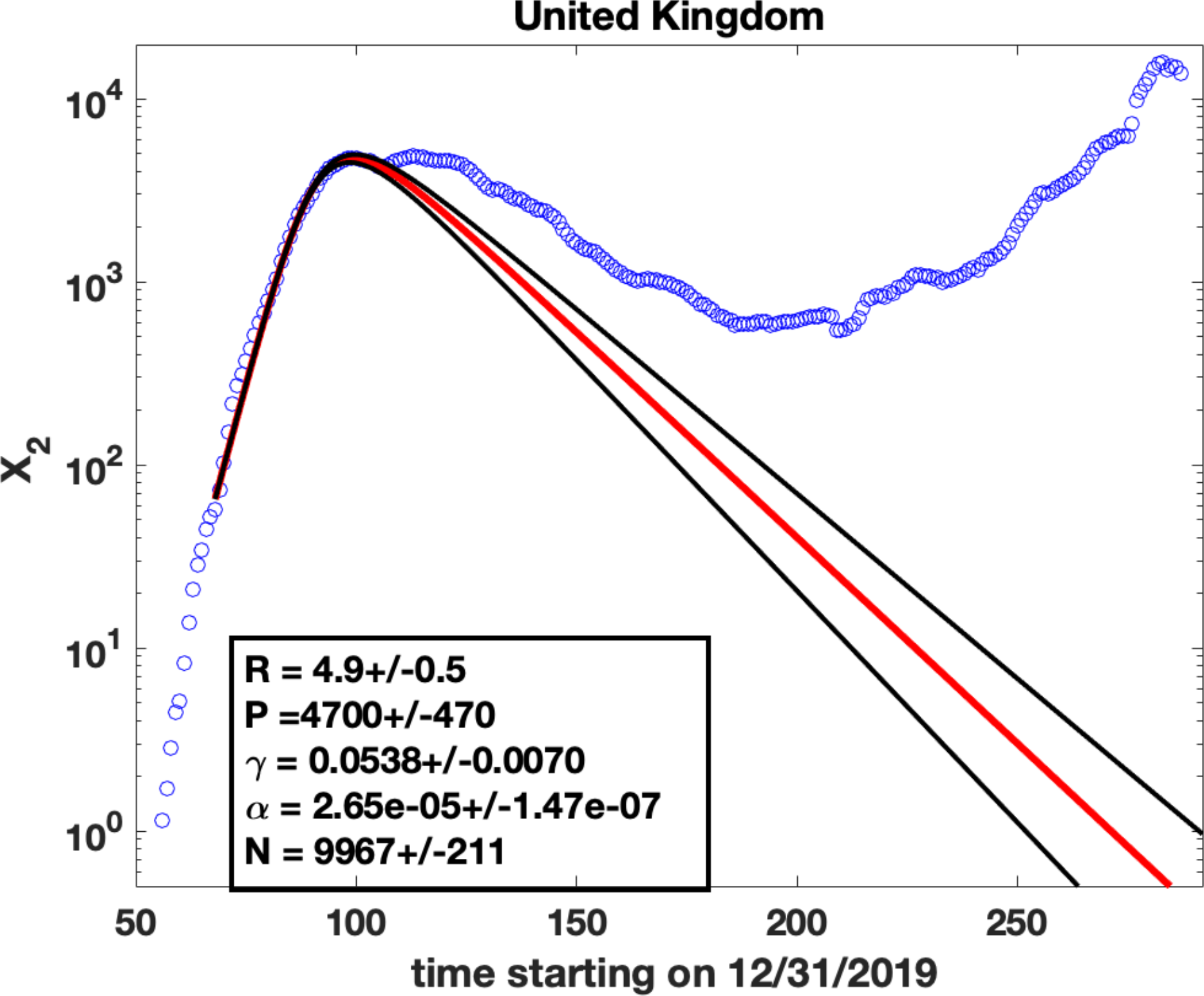
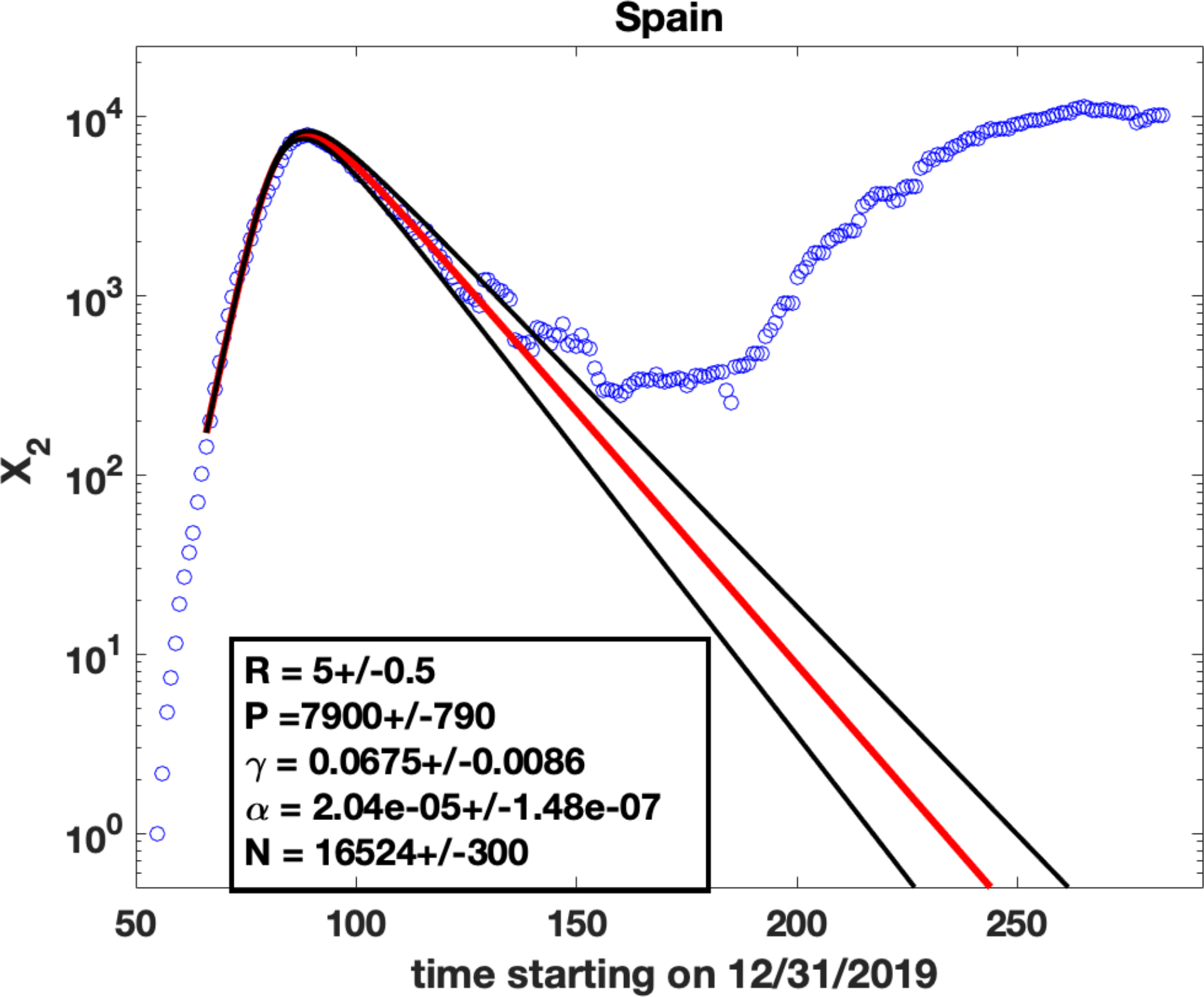
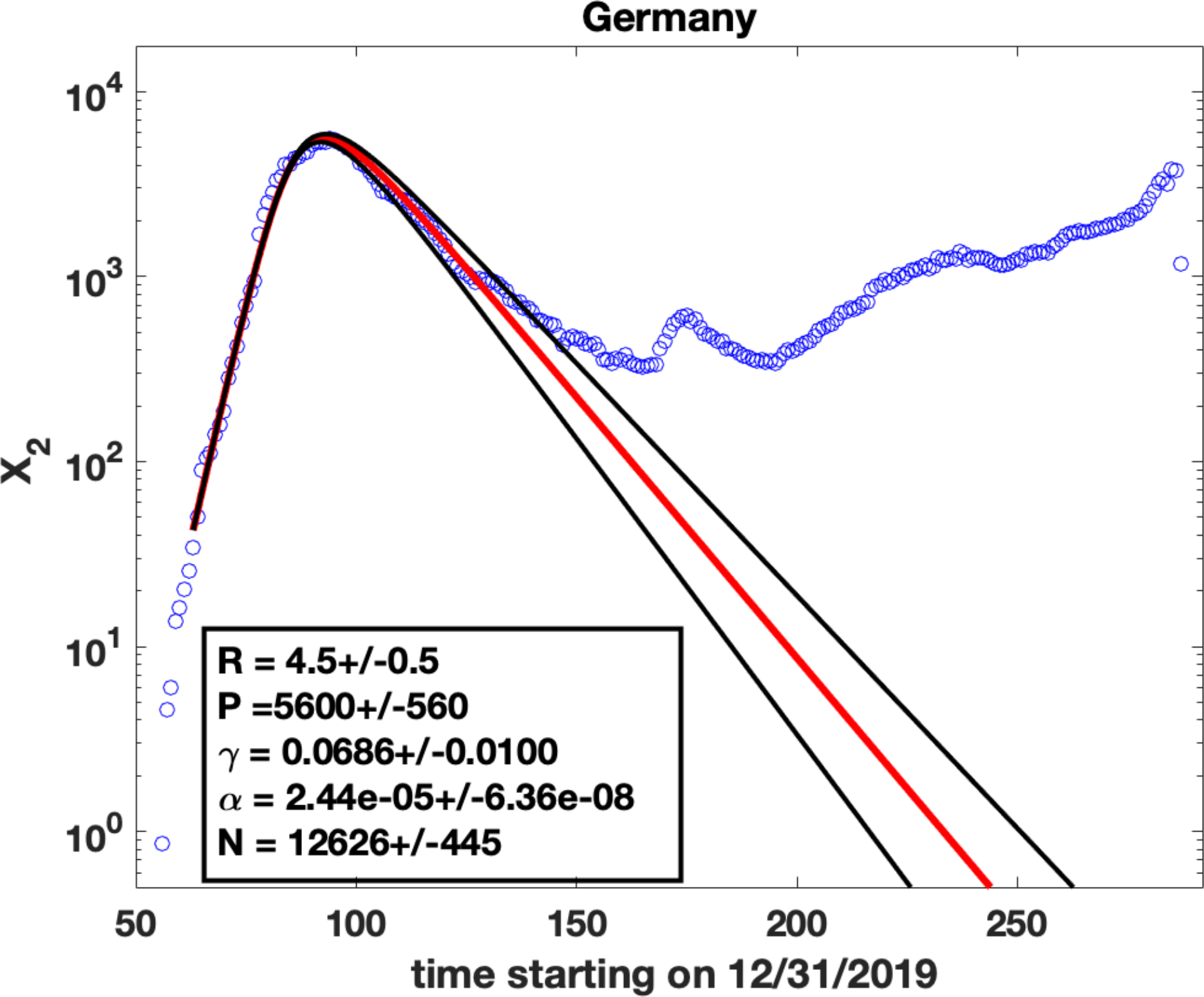
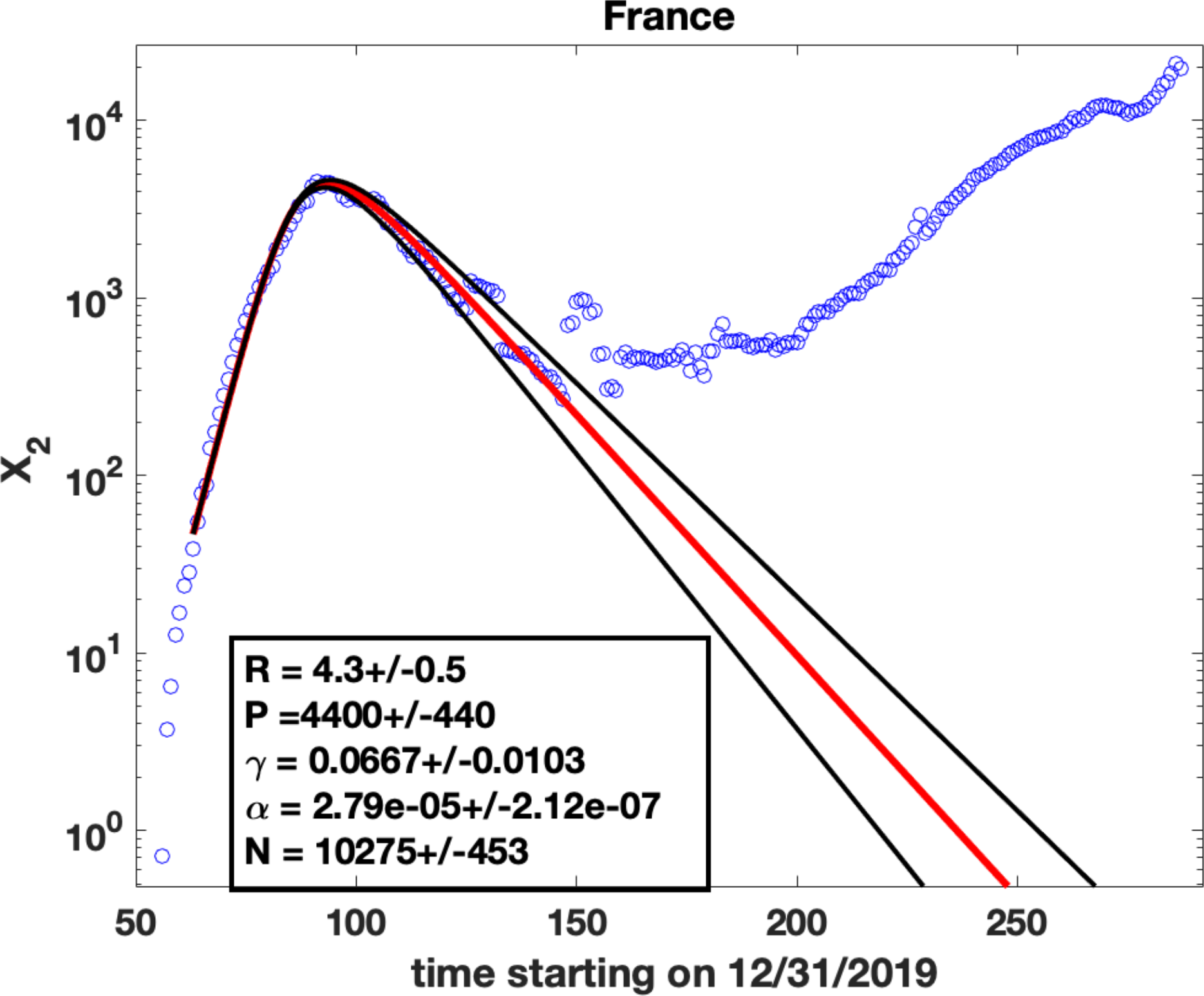
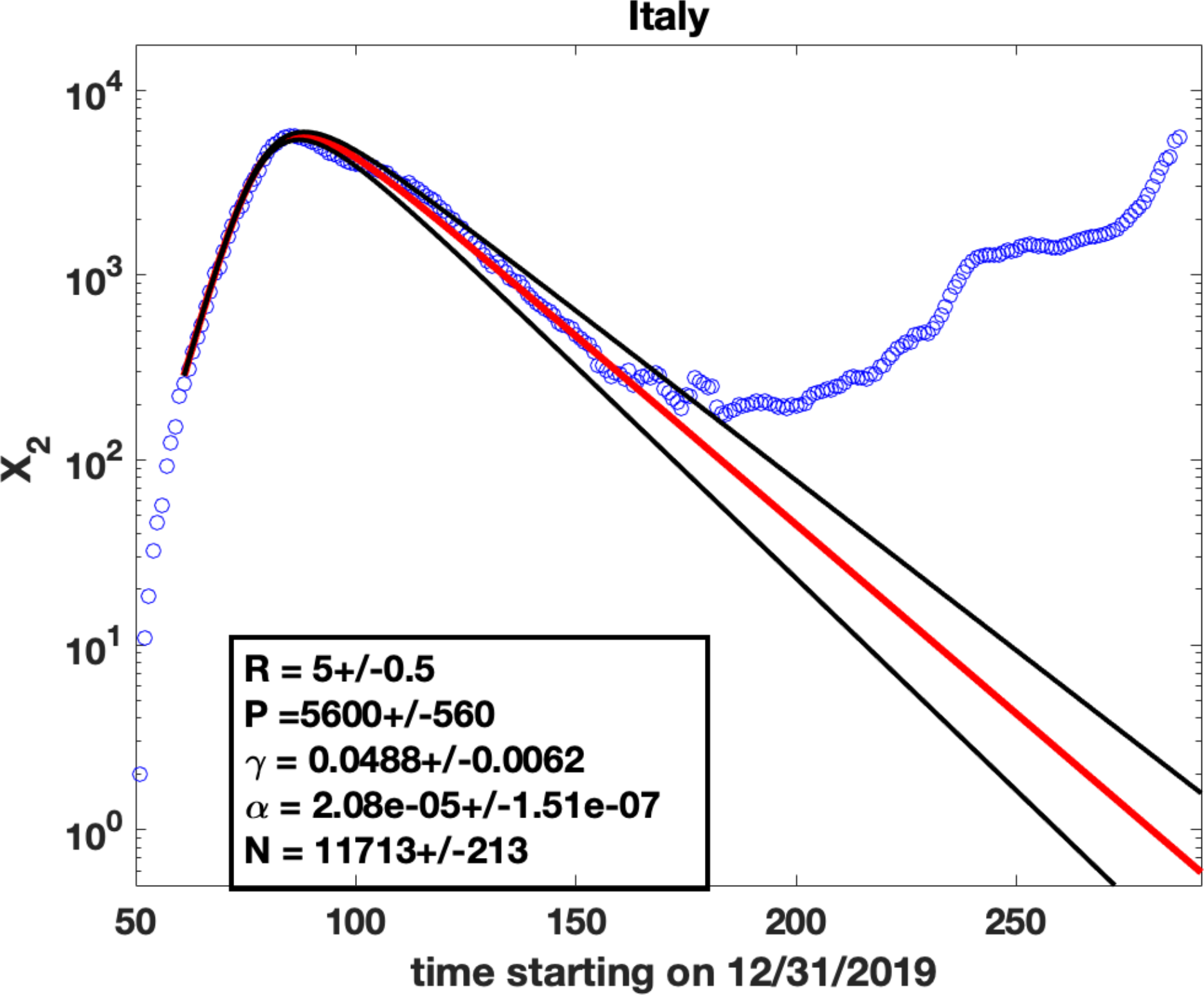
Parameter fits for the number of daily cases: Blue circles are observed data for *X*_2_(*t*), the number of cases per day. Solid lines are fits obtained by solving (8) and (9) using the ODE solver *ode45* in Matlab. The values of the parameters are shown in the insets and represent the fit shown as the solid red line. The method used for the fits was to find *γ*(*R* − 1) from the exponential rise in *X*_2_ at early times (Appendix A), estimate the peak value *P* of *X*_2_ (which gives the value of *N* using (15)) and find an *R* value that best fits the data (red solid line). The black lines represent solver results for R varying by 0.5 from the best fit value.

**Figure 3:**
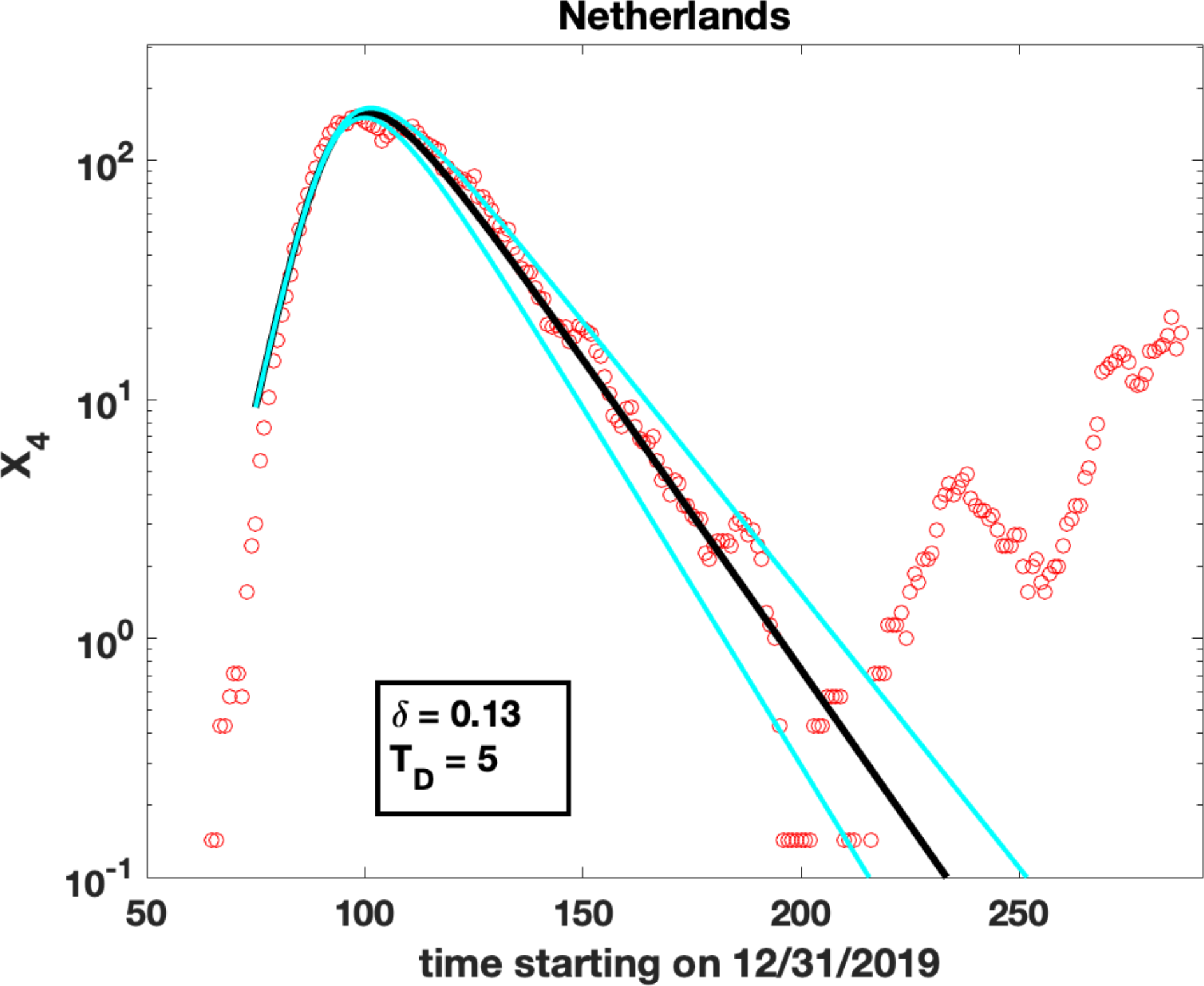
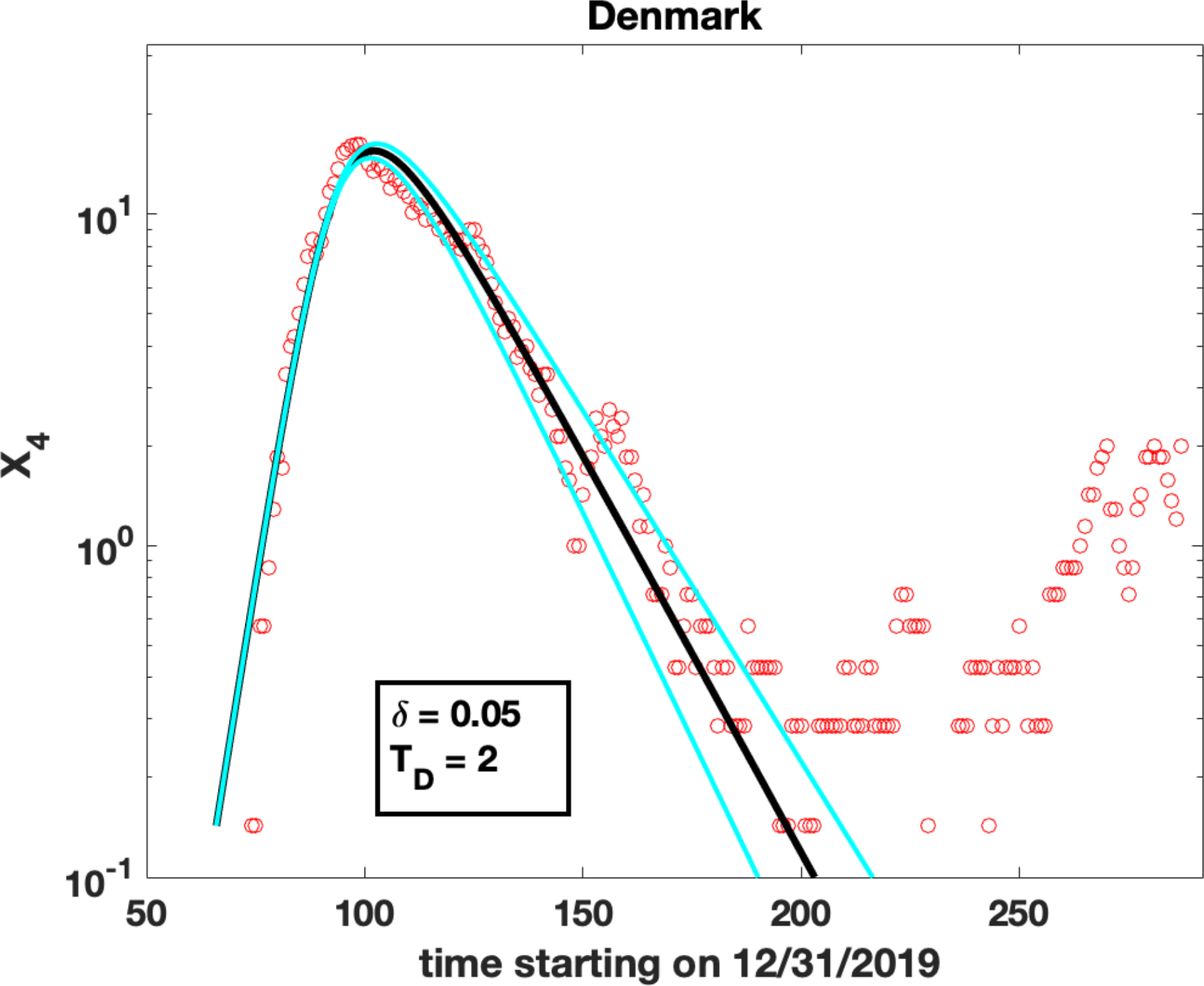
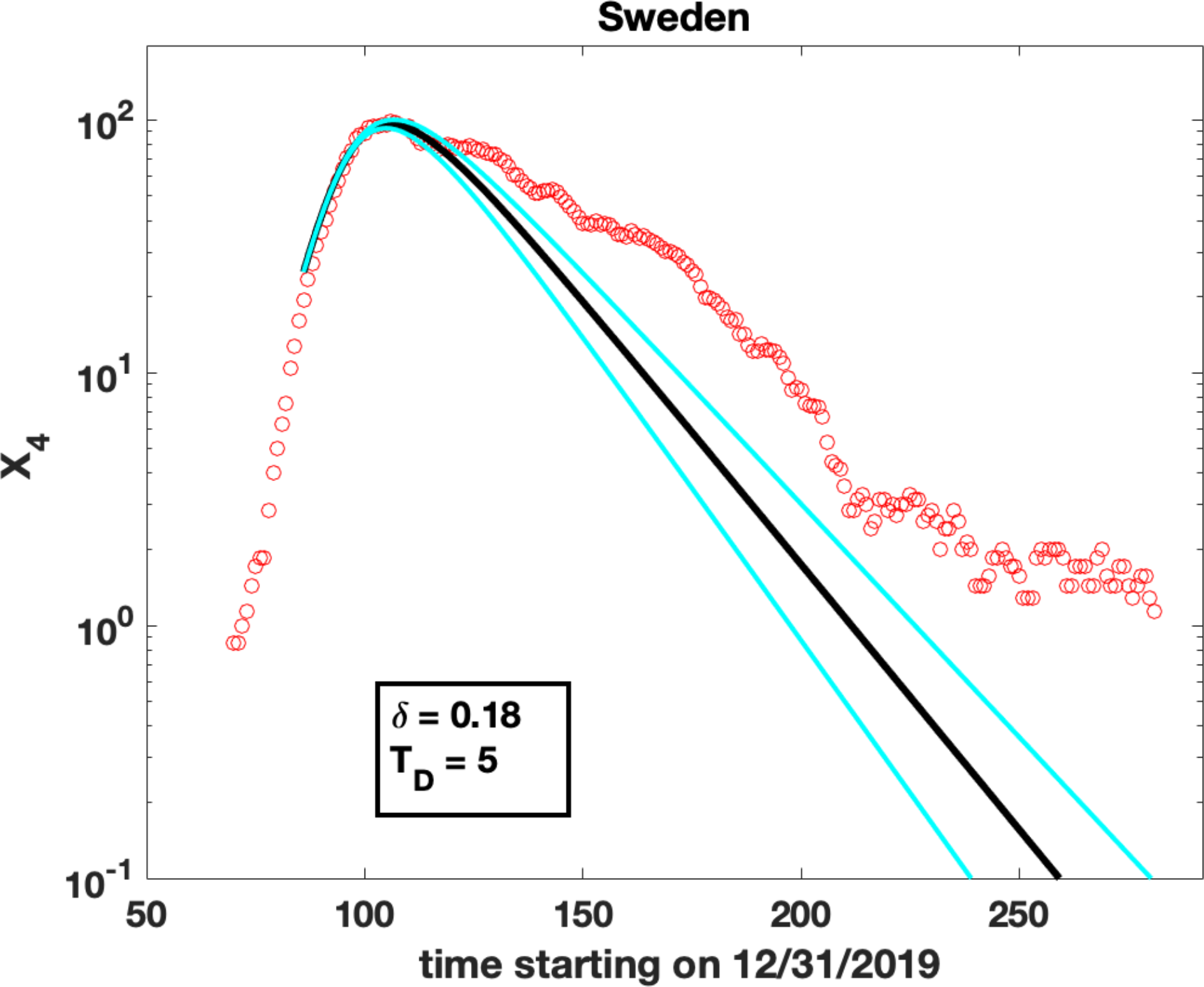
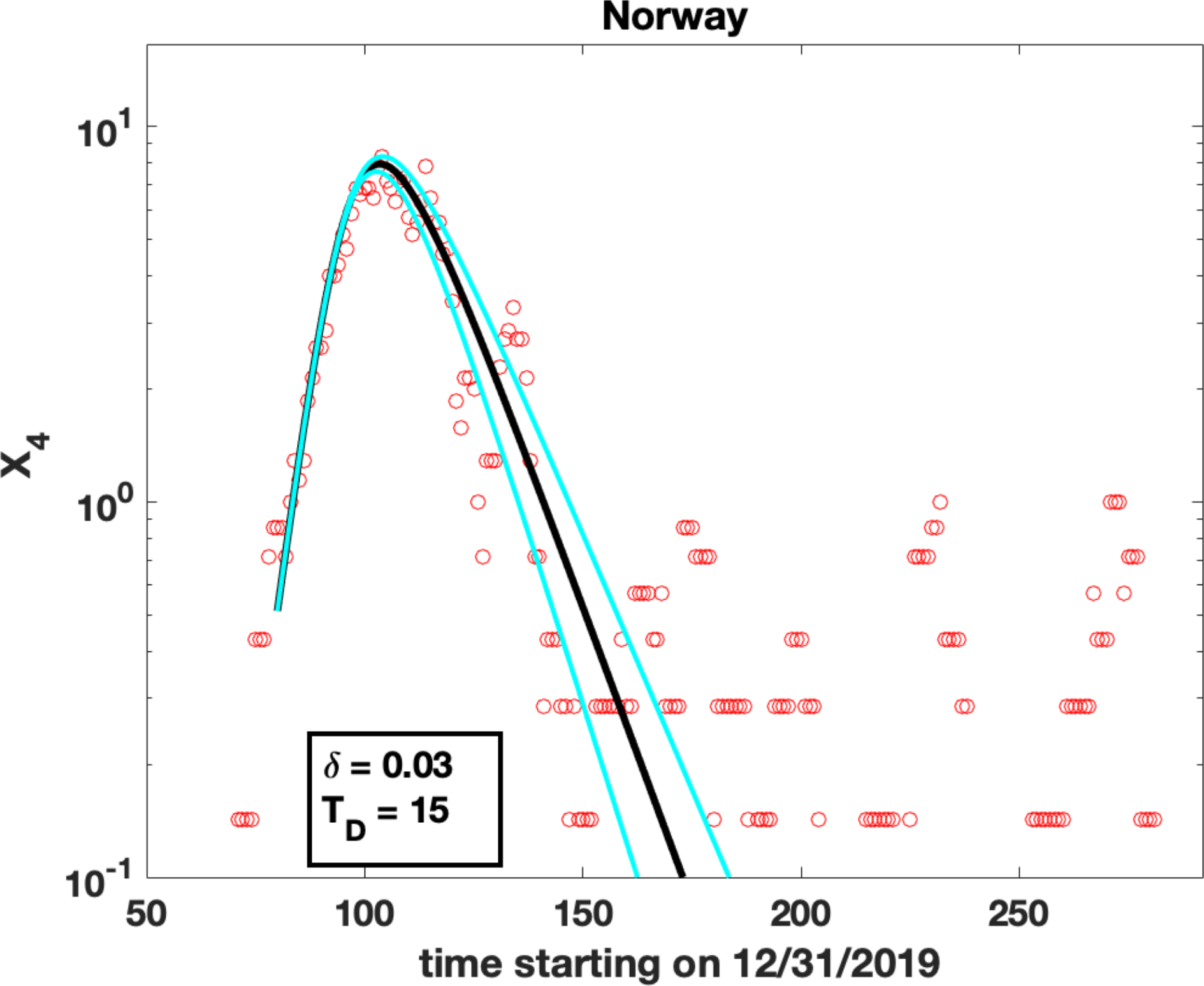
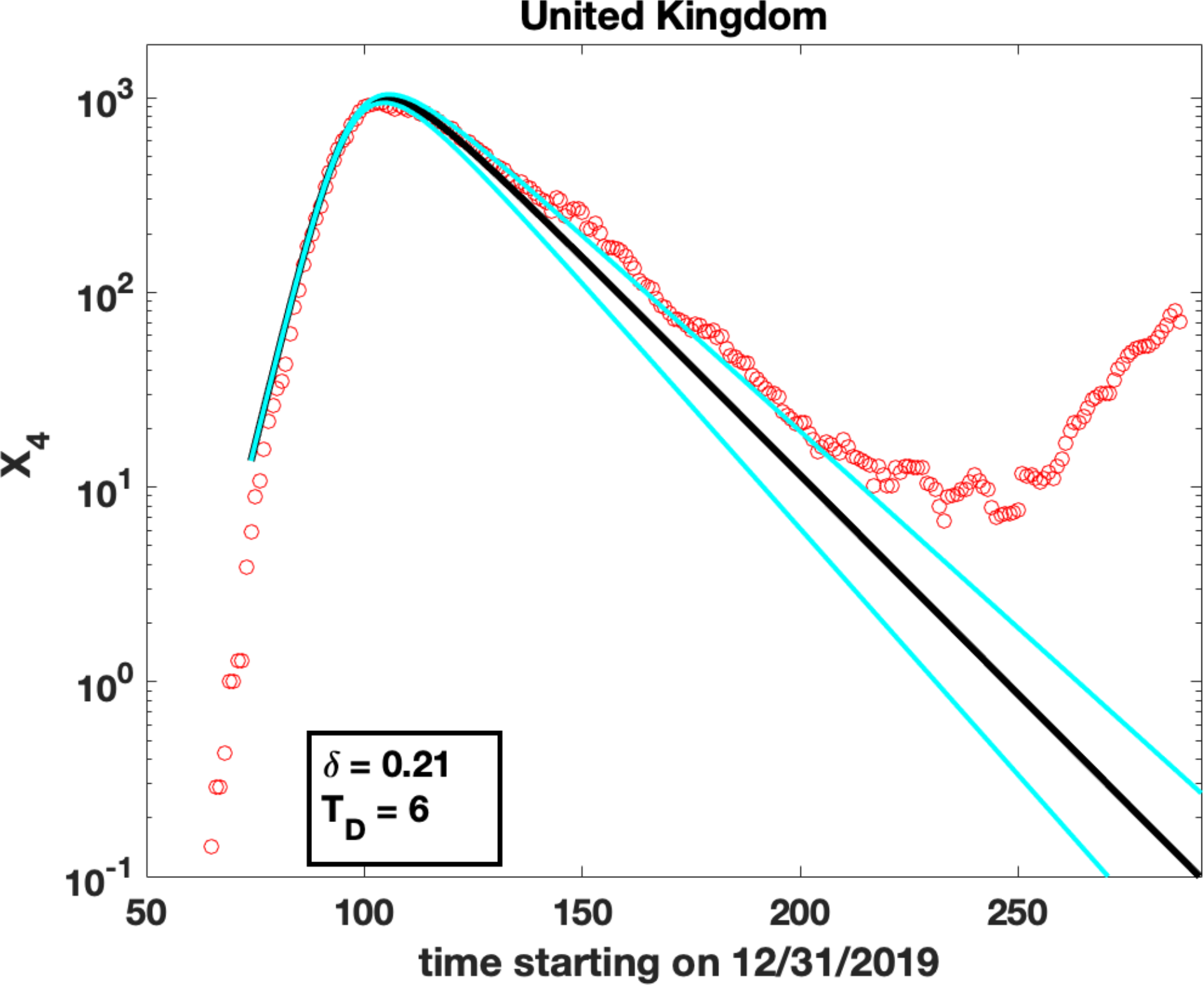
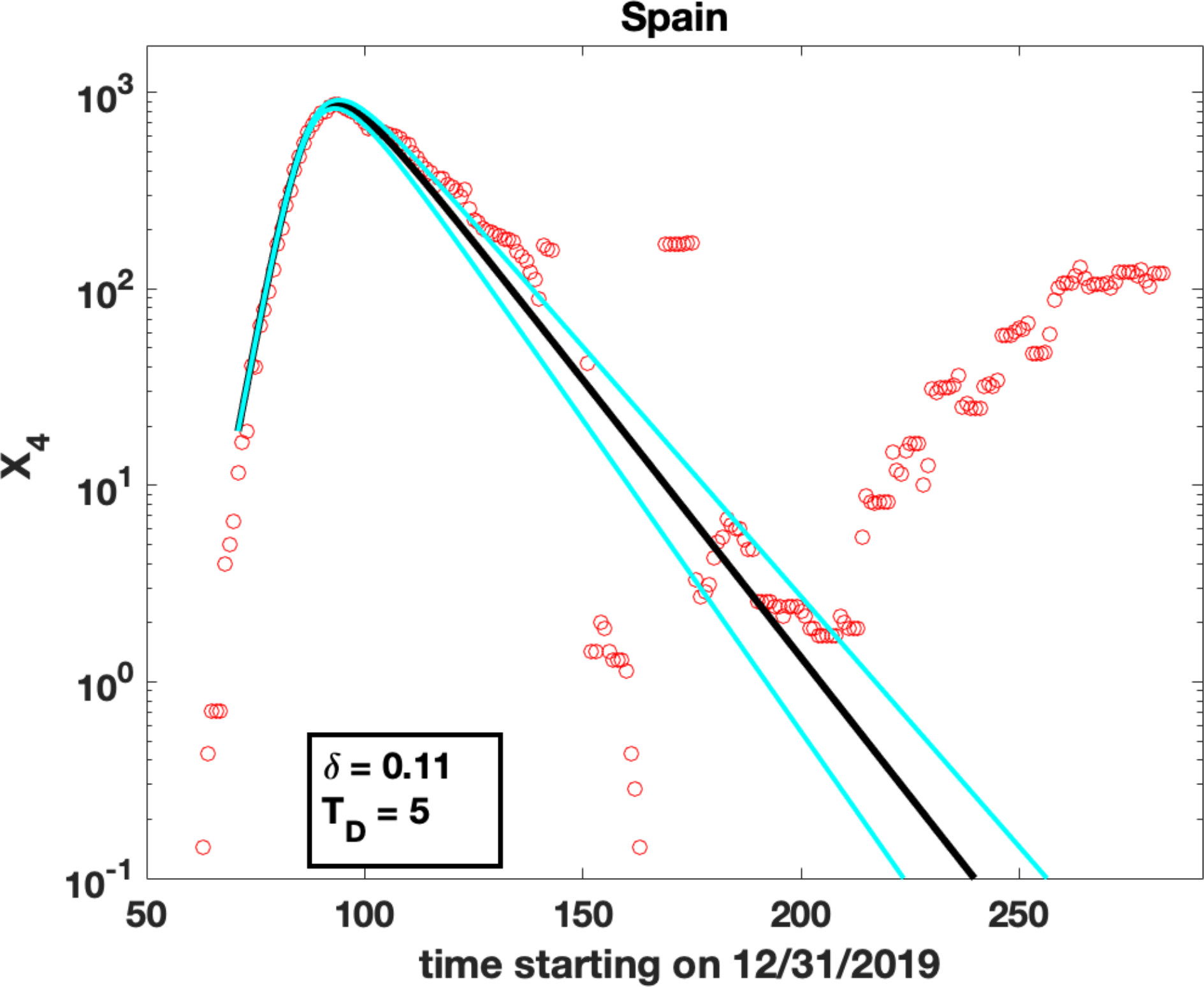
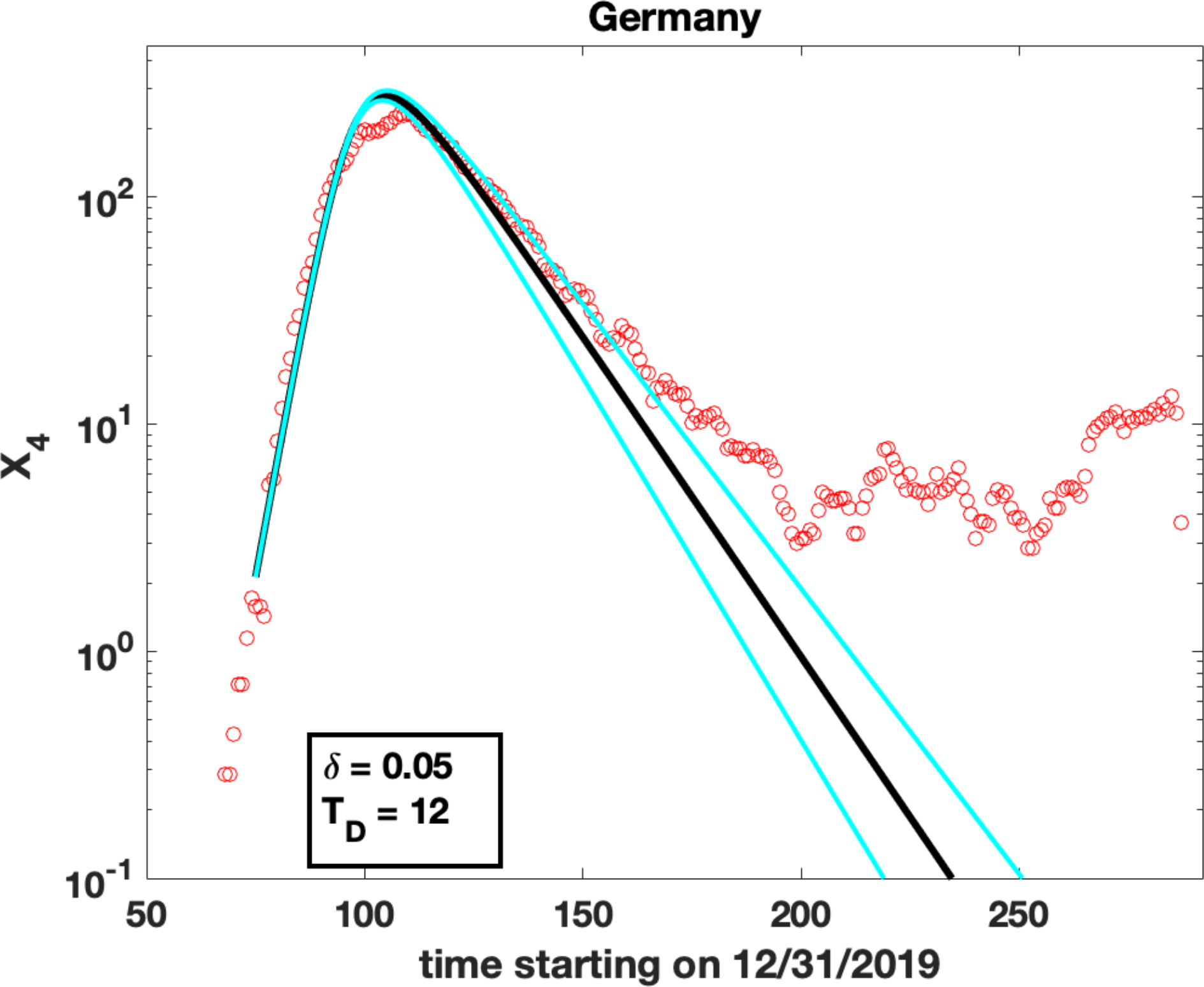
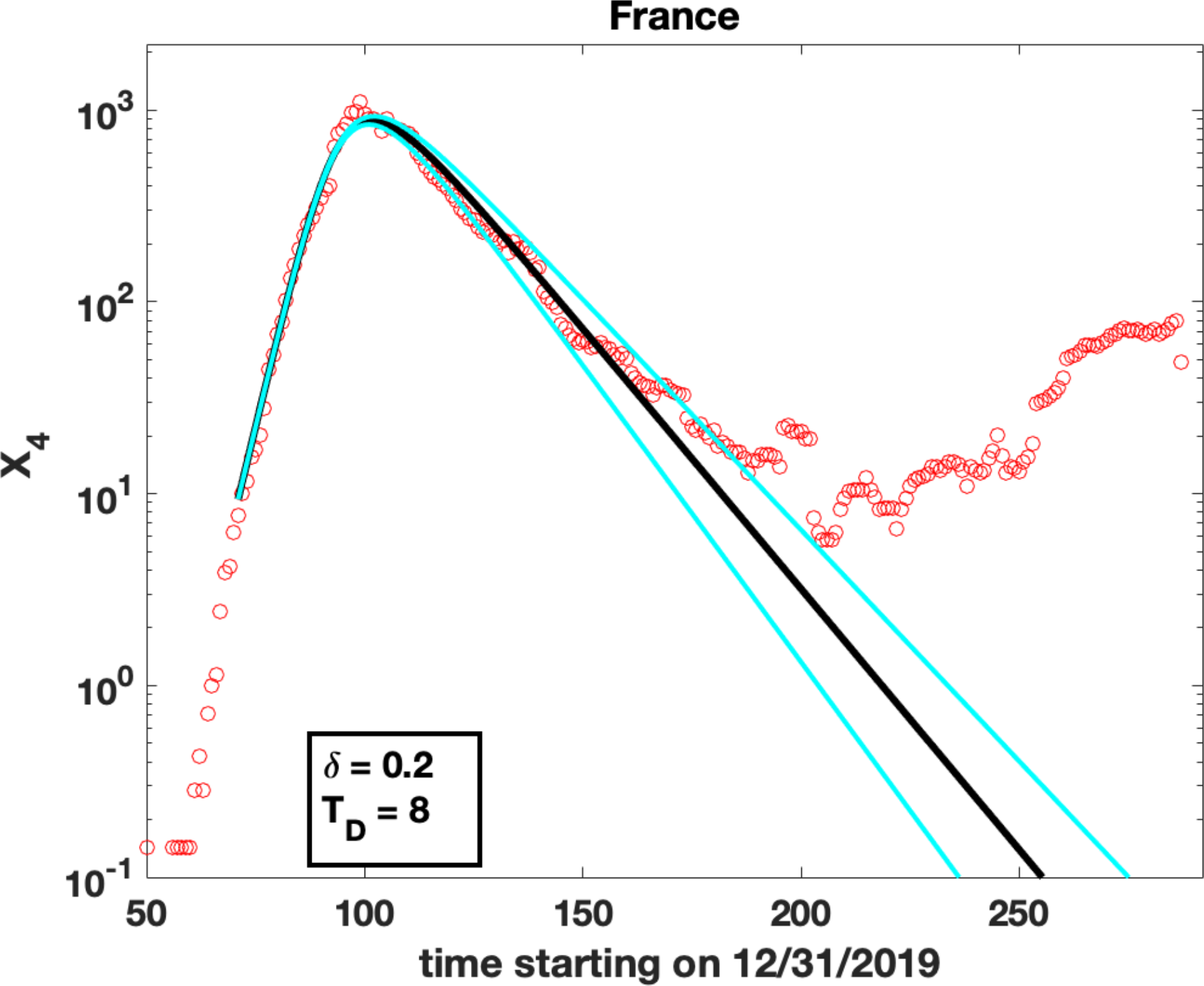
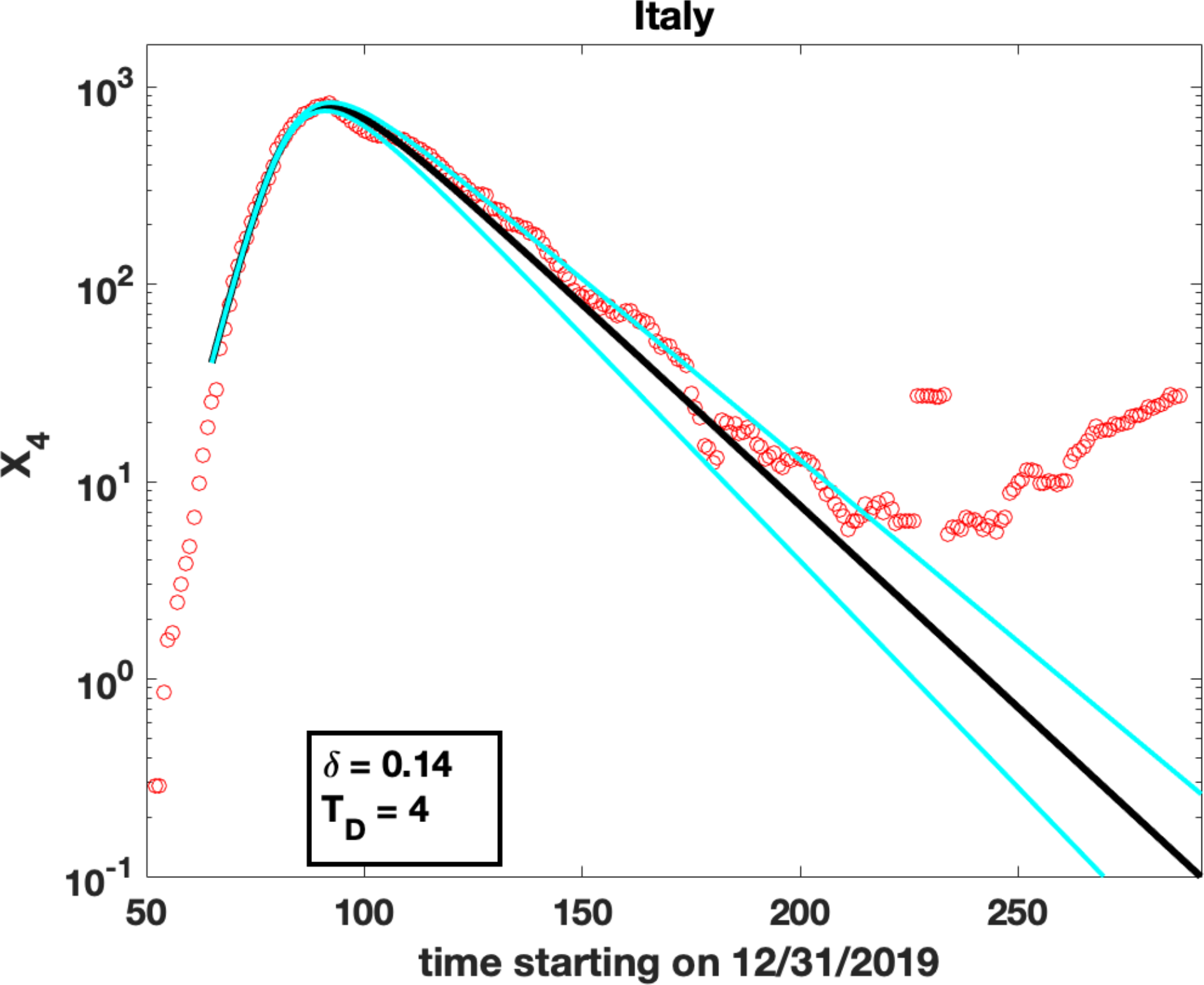
Number of deaths per day and fits for *δ* and *T*_*D*_: Red circles are observed data for *X*_4_(*t*), the number of deaths per day. The solid lines were obtained by shifting the fits for *X*_2_(t) in Figure 2 forward in time by an amount *T*_*D*_ and scaling the results by *δ* (see eq. 5a).

In contrast, for the number of daily deaths *X*_4_ (Figure 3 a-i), there were significantly smaller increases past the peak for all countries, suggesting that measures to save lives worked better than measures to contain the number of infections. This is likely due to the efficiency of the health care systems in these countries and the competence and professionalism of health care workers.

### Results for ***T***_***D***_, ***δ, T***_***L***_, the time delay between case identification and death and the time interval ***T***_***R***_ between transmissions while infective

The fraction *δ* of identified cases who died after a time interval *T*_*D*_ (Figure 4a, Table A) also shows significant variation by country. Norway, Germany and Denmark have the smallest values: *δ* = 0.03, 0.050 and 0.050 respectively, and the UK, France and Sweden have the highest: *δ* = 0.21, 0.20 and 0.18 respectively. Assuming that most of the deaths occurred in hospitals, the average time *T*_*D*_ from case identification to death for those who died was highest (12, 15 days) for Germany and Norway respectively and lowest (2, 4 days) for Denmark and Italy respectively (Figure 4b, Table A).

**Figure 4:**
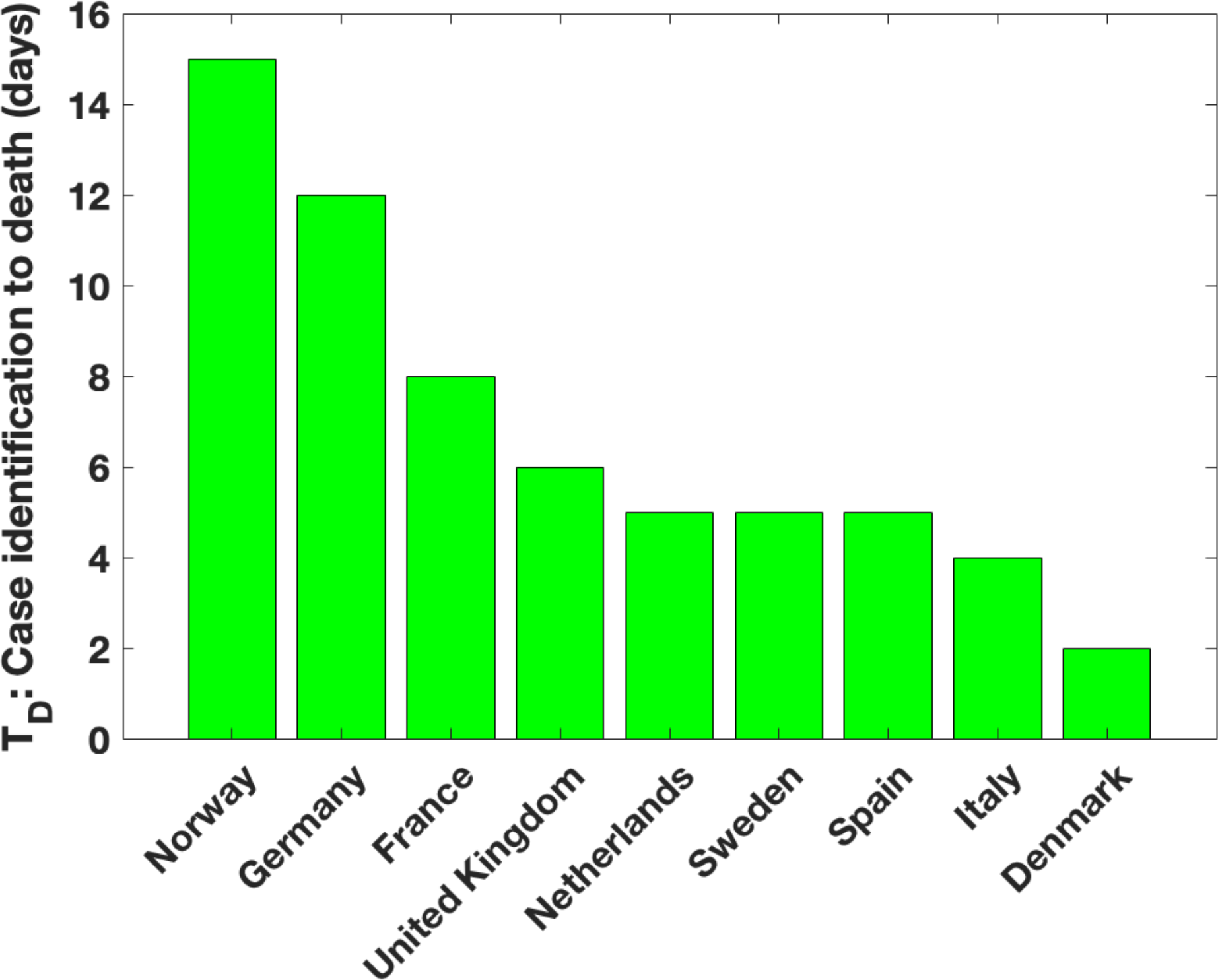
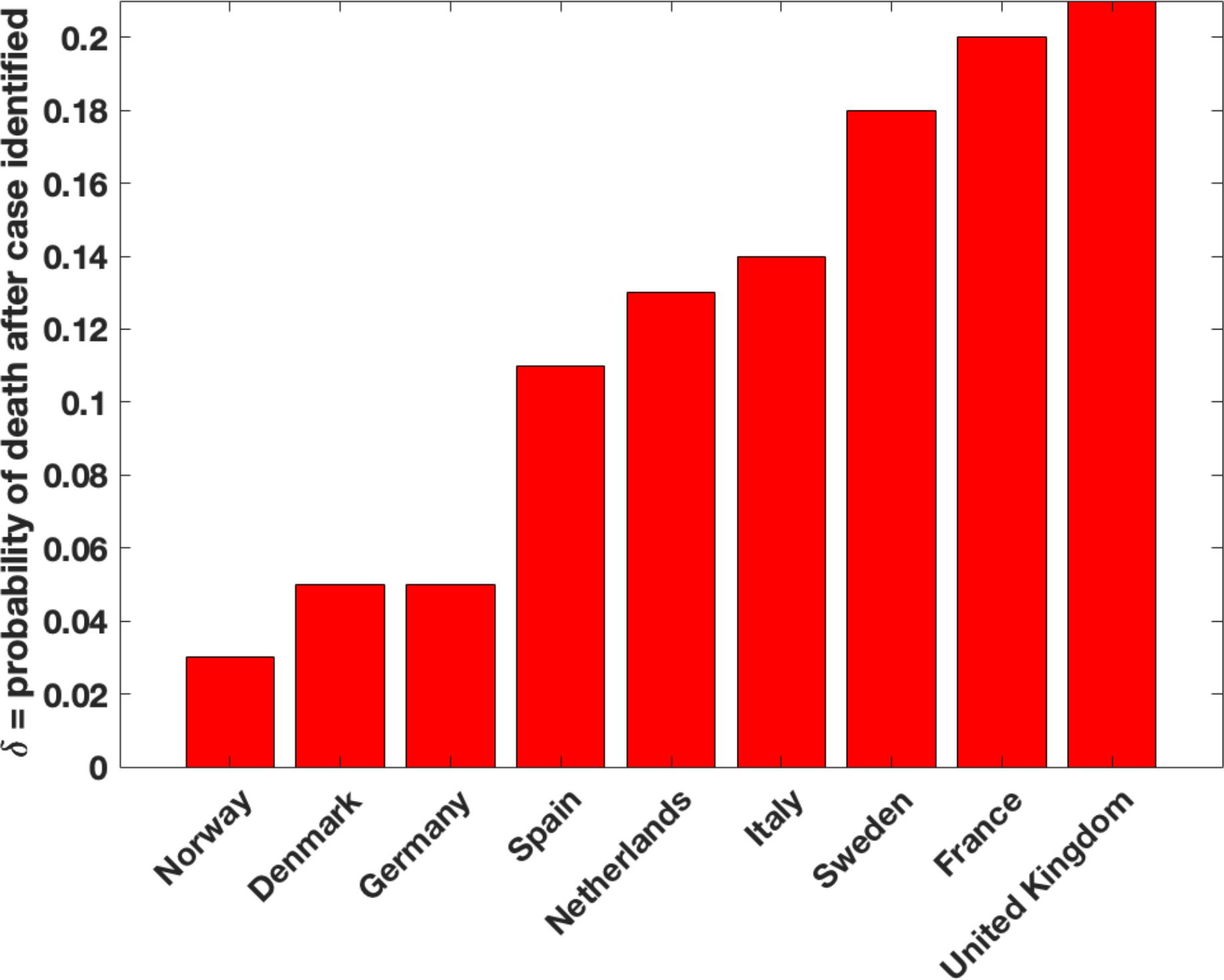
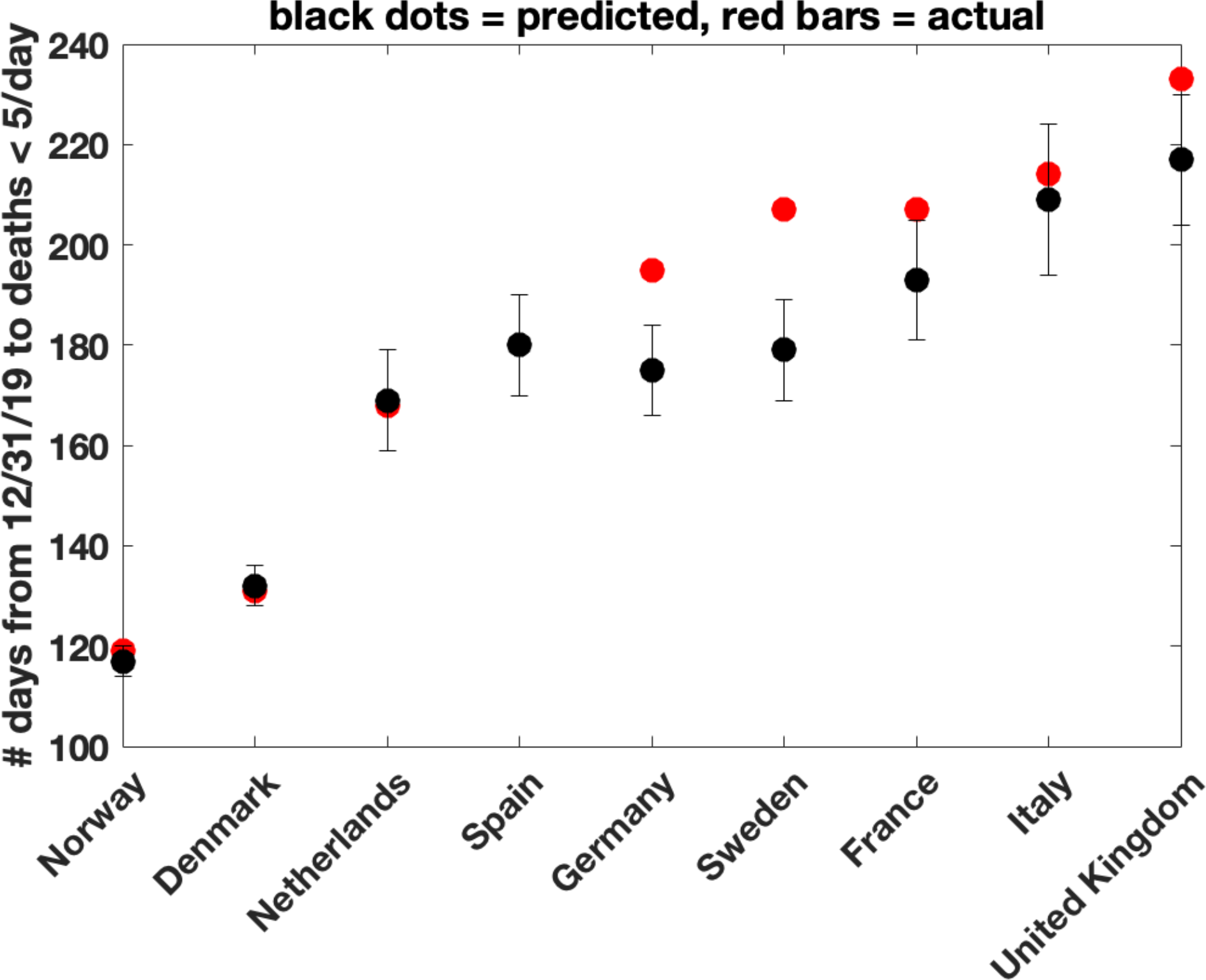
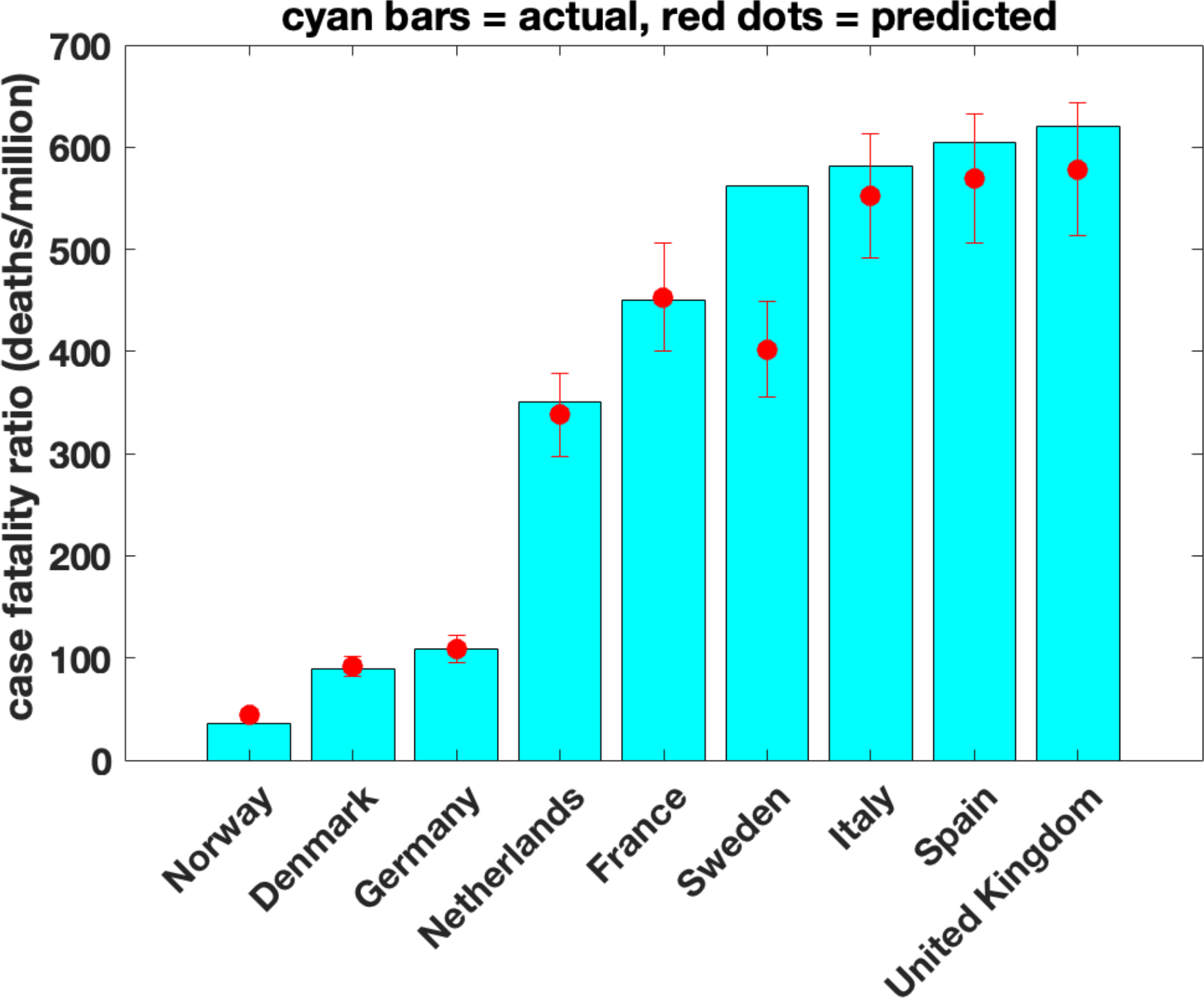
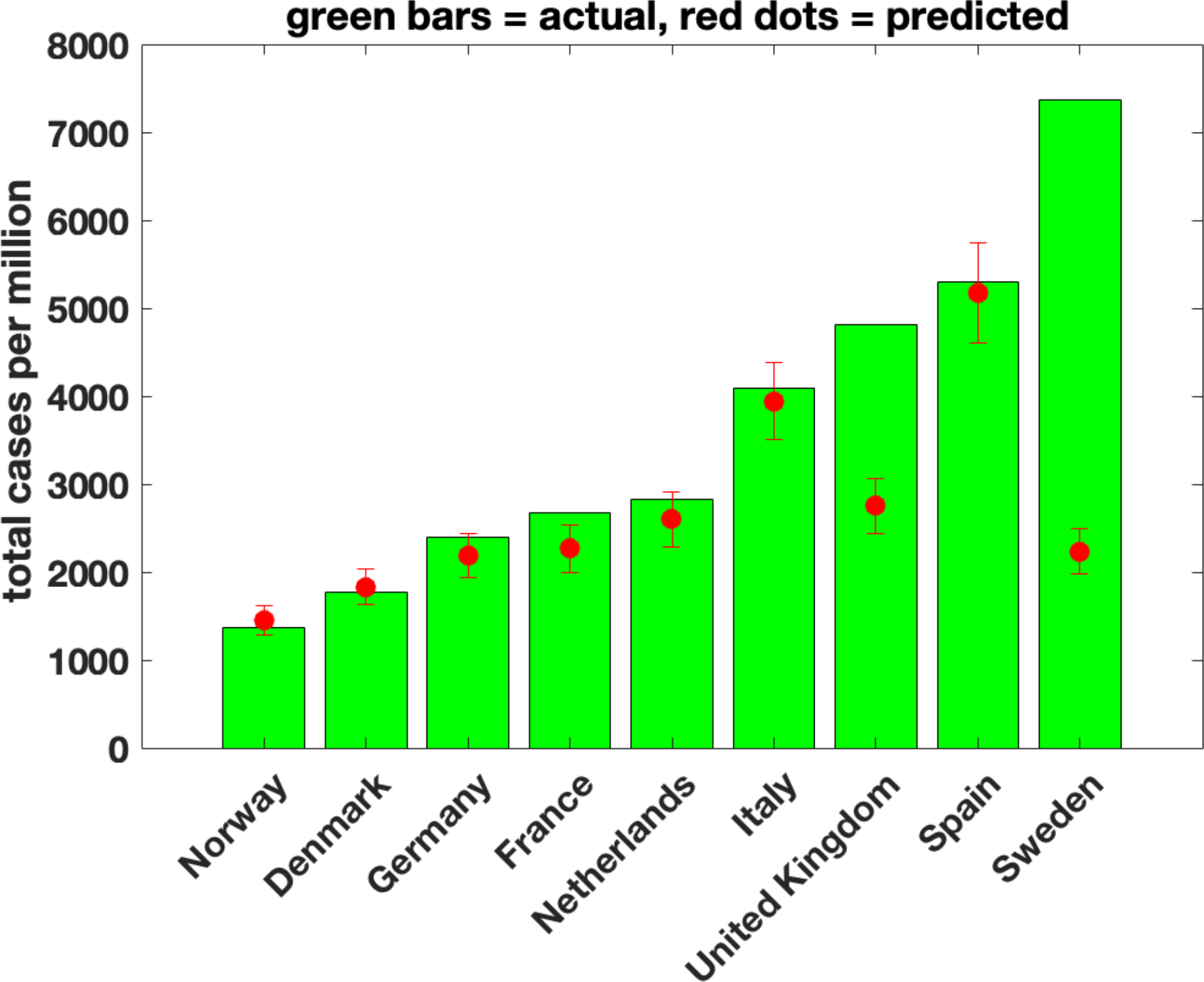
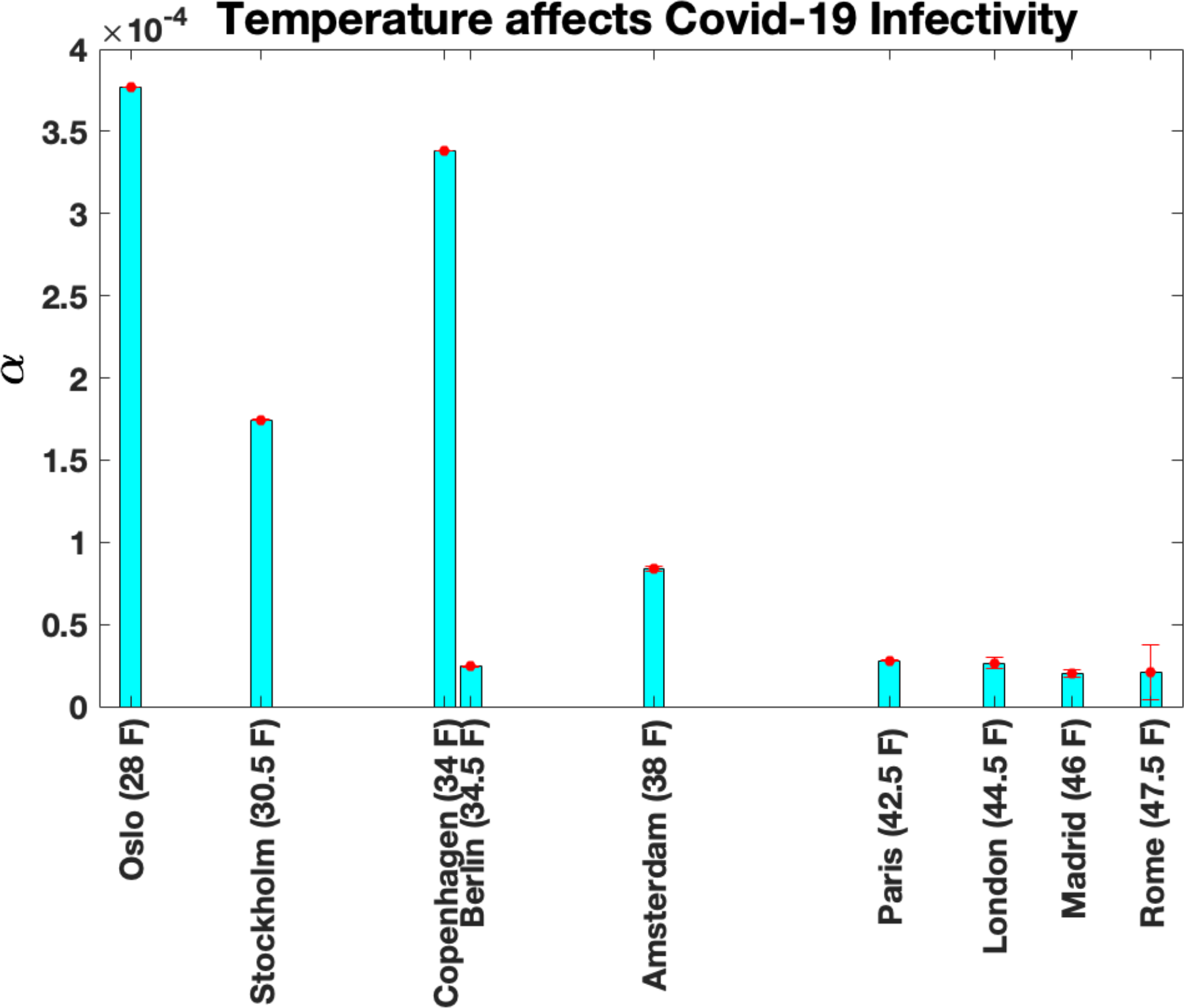
Predictions from fits. **(a)** Best fits for *T*_*D*_ (see eq. 5a and Figure 3), the average number of days from case identification to death, if death occurs. Values range from a high of 15 days for Norway and a low of 4 days for Italy and Denmark. **(b)** Best fits for *δ*, (see eq. 5a and Figure 3), the fraction of identified cases that died an average of *T*_*D*_ days after case was recorded. Values range from low values 0.032, 0.05 and 0.05 for Norway, Denmark and Germany to high values 0.21, 0.20 and 0.20 for UK, Sweden and France. **(c)** Predicted and actual number of days from 12/31/2019 to when number of daily deaths < 5. Note that for all countries, our predictions are very close to the actual results. This is also evident from the extrapolation of the fits past the peak in Figure 3. This suggests that measures to limit fatalities were just as effective after the peak as before. **(d)** Predicted and actual case fatality ratios (number of deaths per million) from start of the pandemic to when daily deaths < 5. Actual values for the case fatality ratios range from lows of 18 and 76 deaths/million population for Norway and Denmark and highs of 617, 604 and 579 deaths/million population for UK, Spain and Italy. Note that the predicted values from the fits are in good agreement with the actual values for all countries, suggesting consistent medical care throughout the pandemic. **(e)** Predicted and actual cases per million population from start of the pandemic to when daily deaths < 5. Note that now, the predicted numbers are in reasonable agreement for all countries except UK and Sweden, where preventive measures such as partial lockdowns, use of masks and social distancing were not mandated/followed. **(e)** Identification of a possible temperature effect on the infectivity parameter *α*. The x-axis shows the temperature for February 2020 for the principal cities of the countries studied on a linear scale. The results suggest that the infectivity of SARS-CoV-2 decreases with increasing temperature (see also [34]).

It is significant that in each country, the relation between the daily number of cases and the daily deaths is described by just two parameters, *δ* and *T*_*D*_ (Eq. 5a), for the entire duration of the pandemic. This is not required a priori. For example, pressure on resources during a peak period of infectivity might have caused a transient increase in the number of deaths per day relative to the number of recovered per day. In such a situation, a single probability *δ* and a single value of *T*_*D*_ need not have sufficed for the entire epidemic. The fact that there is little evidence of variations in these parameters over the pandemic, to within the quality of the data, suggests that although the effectiveness of life saving measures may differ between countries, these measures seem to be relatively insensitive to changes in the case burden.

The average infective period 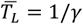 was approximately constant for all countries, with an average value: 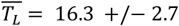 days. The average time between transmissions 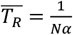 was also remarkably uniform across countries, averaging: 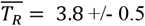 days. Finally, the average number of transmissions while infective or 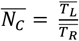, which is also the average value of *R*, varied only in a narrow range for all countries, averaging: 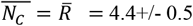.

### The duration of the pandemic, the case fatality ratio (deaths/million) and cases/million

We define the end of the pandemic as the day when daily deaths becomes less than 5 for the first time after first peaking. The duration of the pandemic in each country is shown in Figure 4c as the number of days from 12/31/2019 till the day that daily deaths became < 5. With this definition, the predicted and actual dates are (Table A): Netherlands: (6/18 +/- 10 days, 6/17), Denmark: (5/11 +/- 4 days, 5/10);, Sweden: (6/28 +/- 10 days, 7/26), Norway: (4/26 +/- 3 days, 4/28), UK: (8/5 +/-13 days, 8/21), Spain: (6/29 +/-10 days, 6/29), Germany: (6/24 +/- 9 days, 7/14), France: (7/12 +/-12 days, 7/19), Italy: (7/28 +/- 15 days 8/2). Note that the model predictions for pandemic duration are in quite good agreement with observation.

The measured case fatality ratio or the total number of deaths per million population (Table A) ranged from low values of 36 and 89 for Norway and Denmark, to high values of 621, 604 and 582, 562 for the UK, Spain, Italy and Sweden respectively. The model predicted values for this quantity are in good agreement with the actual values (Figure 4d).

The measured total number of cases per million population (Table A) ranged from low values of 1367 and 1774 for Norway and Denmark to high values of 7375, 5293, 4806 and 4092 for Sweden, Spain, Sweden, the UK and Italy respectively. In UK and Spain, the model predictions for this quantity were in sharp disagreement with the actual numbers (Figure 4e), likely the result of a significant relaxation of containment measures after the peak in the daily cases was reached.

### SARS-Cov-2 may transmit less effectively at higher temperatures

As was also noticed in [34], an interesting finding was a weak “Temperature Effect” on the value of the infectivity parameter *α*. Figure 4f shows the average temperature in February 2020 for the principal cities in each country versus the fitted value of *α*, and suggests that SARS-Cov-2 may transmit less efficiently at higher temperatures. However, this result is far from final and needs further testing and validation because of many confounding factors, such as humidity, use of air conditioning and exhaust fans, crowding, population density etc. that may also have significant effects on *α*.

In summary, the simple extension of the original SIR model [29] that we propose in this paper is able to identify the full set of pandemic parameters and fully characterize all three compartments S, I and R. It would be useful to try to understand what policy measures would result in improved values for some of these parameters. However, it is also important to recognize that some of the parameters cannot be controlled by policy changes, and some that can be will not all respond to the same strategy. For example, *T*_*L*_ can be shortened by increasing the frequency and number of tests, since both these strategies increase the number of people removed from the infecting pool. On the other hand, the probability that an infected individual will die as opposed to recover might be affected by resources such as ventilators and masks. These considerations lead us to ask whether the degree of correlation between the parameters of the model, and features of the pandemic such as duration and deaths per million population, can account for the wide intercountry variation in pandemic features. A low degree of correlation for a particular parameter would indicate that it had little influence on variation of the feature, whereas a high degree of correlation would be informative.

To estimate some dependencies that may have useful policy implications, we computed the Spearman Correlation between the derived parameters *T*_*L*_, N*α*, and *δ* and some metrics of value in determining policy – namely “PD = pandemic duration” and “CFR = case fatality ratio or deaths per million.” We found that *Nα* was not correlated with these metrics (Correlation with PD: −0.20, p-value 0.60; Correlation with CFR: −0.02, p-value 0.98). However, *T*_*L*_ was somewhat correlated (Correlation with PD: 0.64, p-value 0.07; Correlation with CFR: 0.533, p-value 0.15) and *δ* was highly correlated (Correlation with PD: 0.86, p-value 0.005; Correlation with CFR: 0.76, p-value 0.02). These results suggest that the optimum method to decrease both the duration of the pandemic and the case fatality ratio would be to decrease the infective period *T*_*L*_ and *δ*, which are achievable by early case identification, contact tracing and quarantine (which would reduce *T*_*L*_) and improving quality of care for identified cases (which would reduce *δ*). Whereas these relationships may seem obvious, and indeed have been used as mechanisms to limit the impact of the pandemic in many countries, the fact that they are also readily identifiable in the data is, in our opinion, quite interesting.

The value of *Nα*, the average number of new infections per day, has clear implications for the time it will take a randomized trial to establish therapeutic efficacy. While a statistical analysis is outside the scope of this manuscript, we note that the wide inter-country dispersion in this parameter indicates that the choice of population could have a pronounced effect on the time to establish efficacy, and it might therefore be best to select a test population deliberately from a country or countries with large and similar values of this parameter, rather than mix populations from different countries with no consideration of the average rate of infectivity.

## Supporting information

Table A excel version

## Data Availability

The data used in this paper were all derived from public sources. Links to these data are included in the paper. The Matlab codes used to analyze the data along with all data files will be provided on request -.

https://ourworldindata.org/coronavirus-source-data

https://ourworldindata.org/coronavirus-testing

## Funding and Acknowledgments

GB was partly supported by grants from M2GEN/ORIEN, DoD/ KRCP (KC180159) and NIH/NCI (1R01CA243547-01A1). He thanks Professors Pablo Tamayo and Jill Mesirov for their kind hospitality at UC San Diego during his sabbatical year 2019-2020 when this work was done. GB thanks Kevin Raines, Sebastian Doniach, Alain Billoire and Carl Dietrich Foerster for many helpful discussions and suggestions.

## Declarations

### Ethics approval

Not applicable

### Consent to participate

Not applicable

### Consent for publication

Not applicable

### Availability of data and material

The data used in this paper were all derived from public sources. Links to these data are included in the paper.

### Code availability

The Matlab codes used to analyze the data along with all data files will be provided on request. Please send an email to if you want these codes.

### Author Contributions

GB: Idea development, analysis, manuscript.

CD: Idea development, analysis, manuscript.

## Supplementary Figure Legend

**Supplementary Figure S1:**
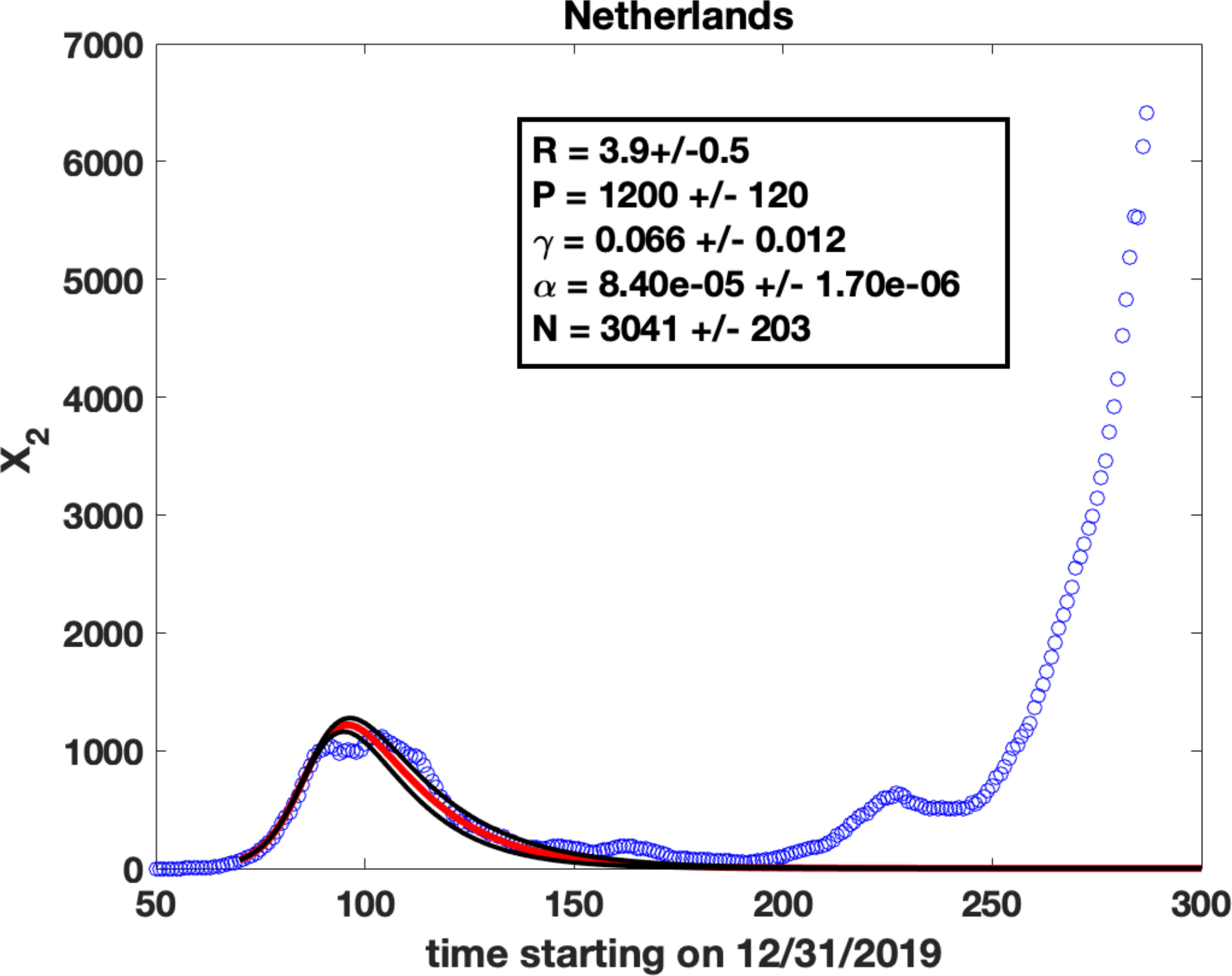
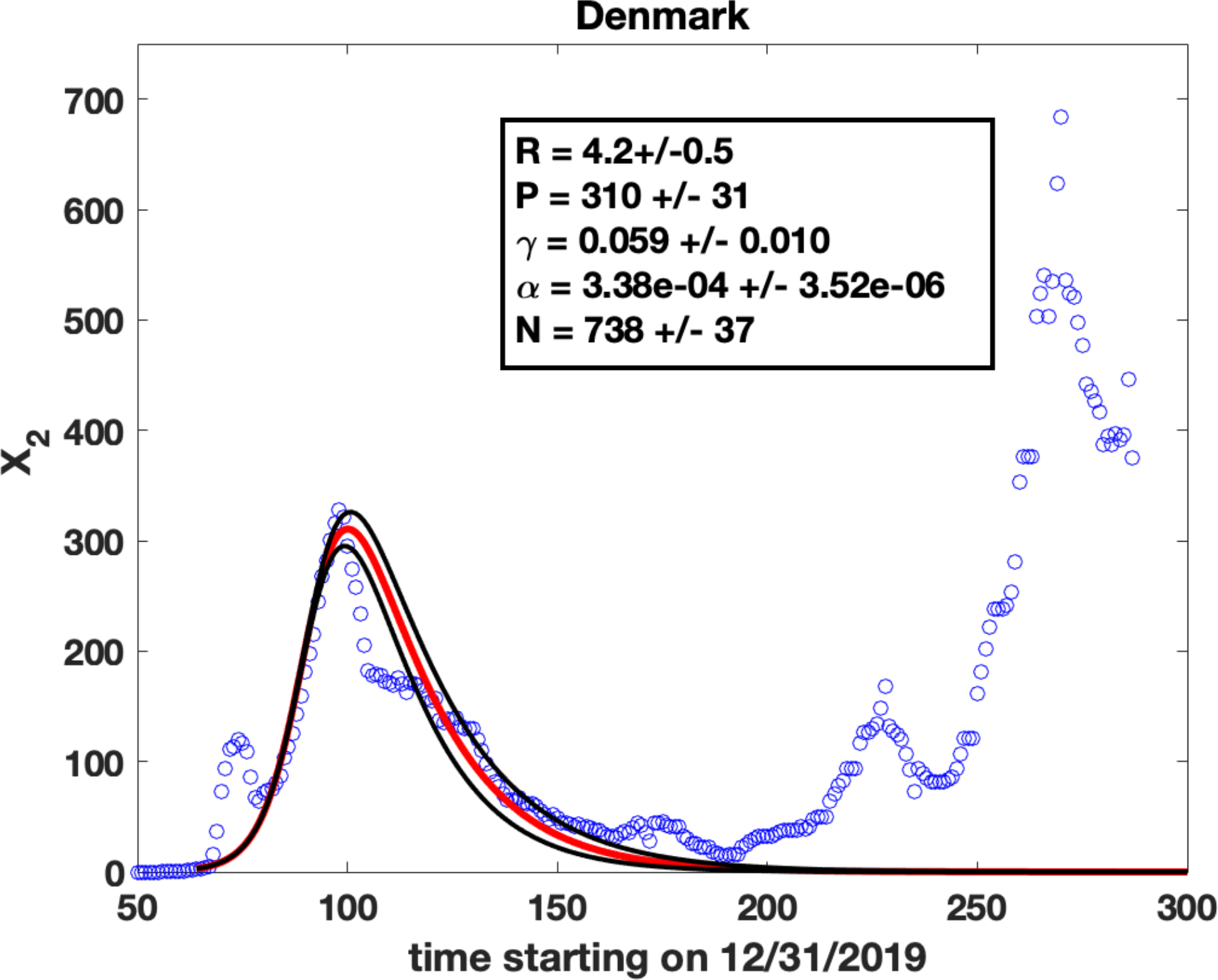
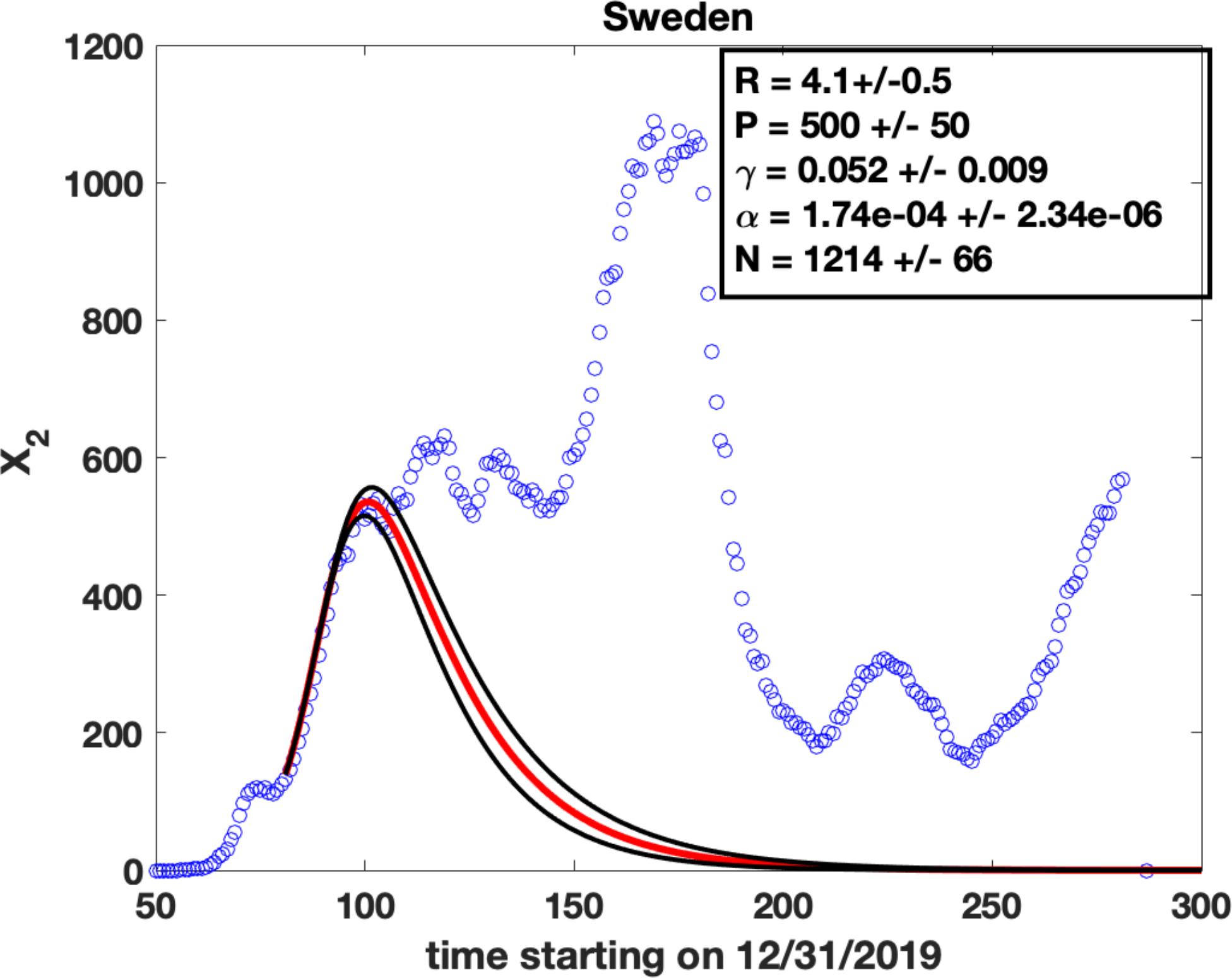
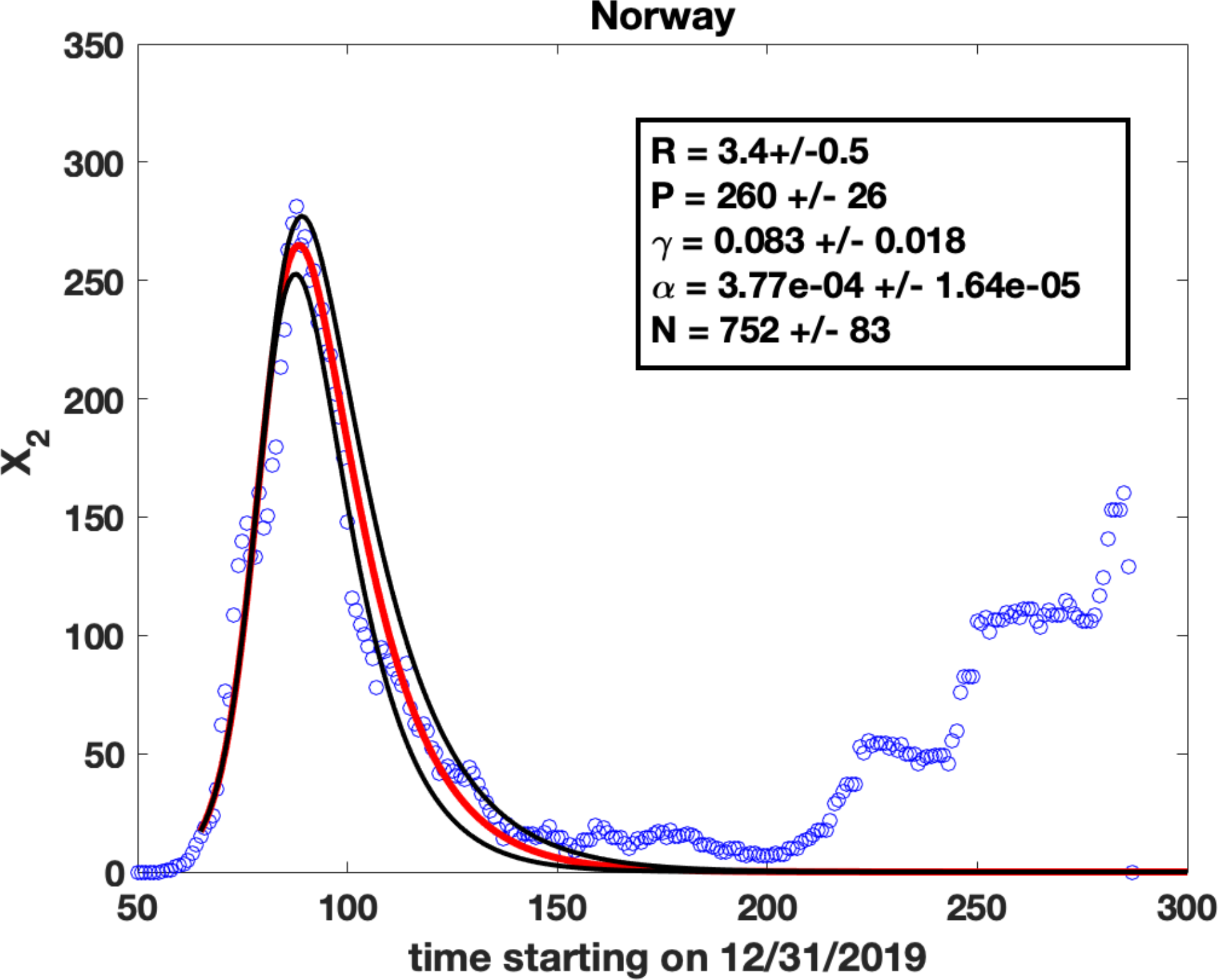
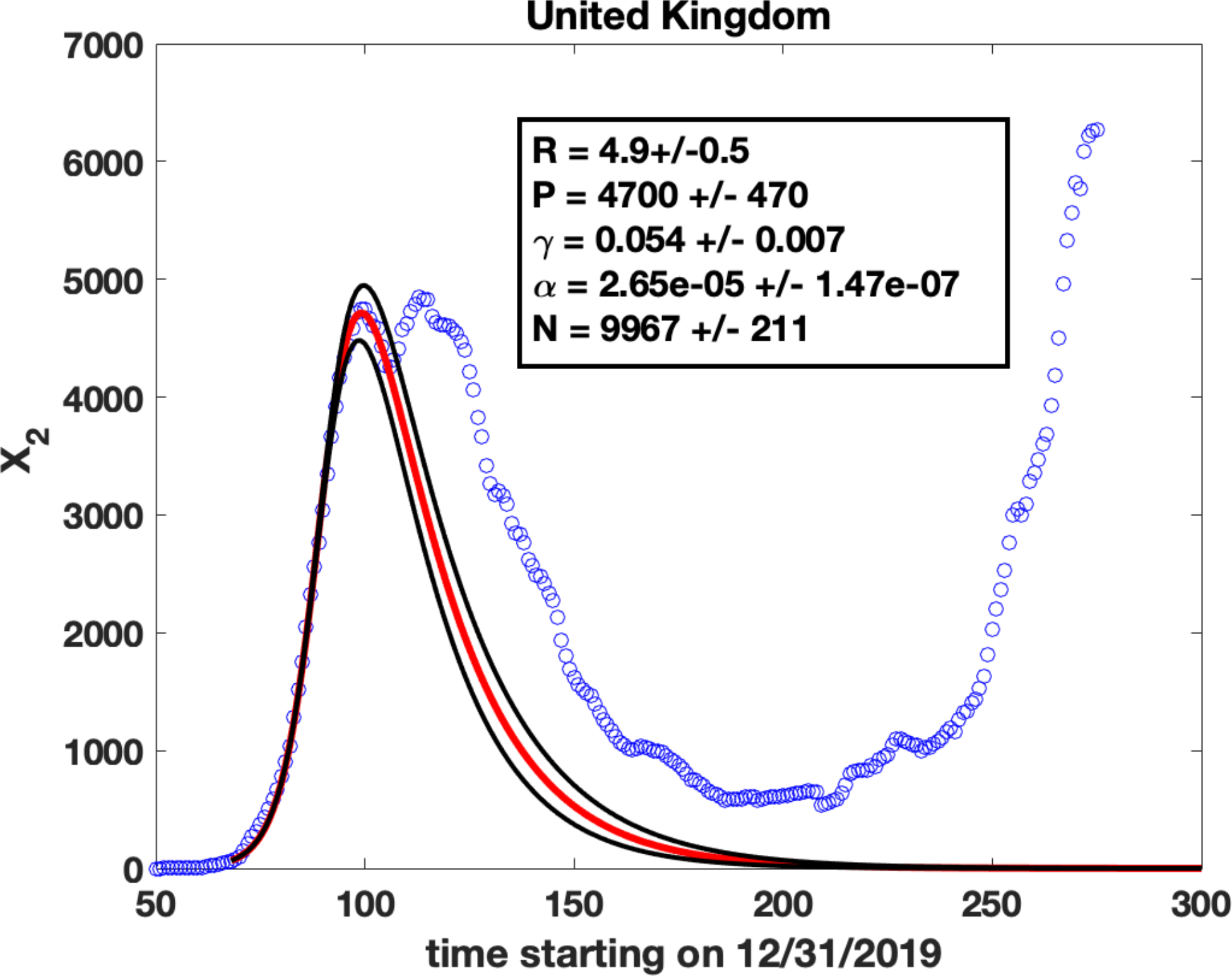
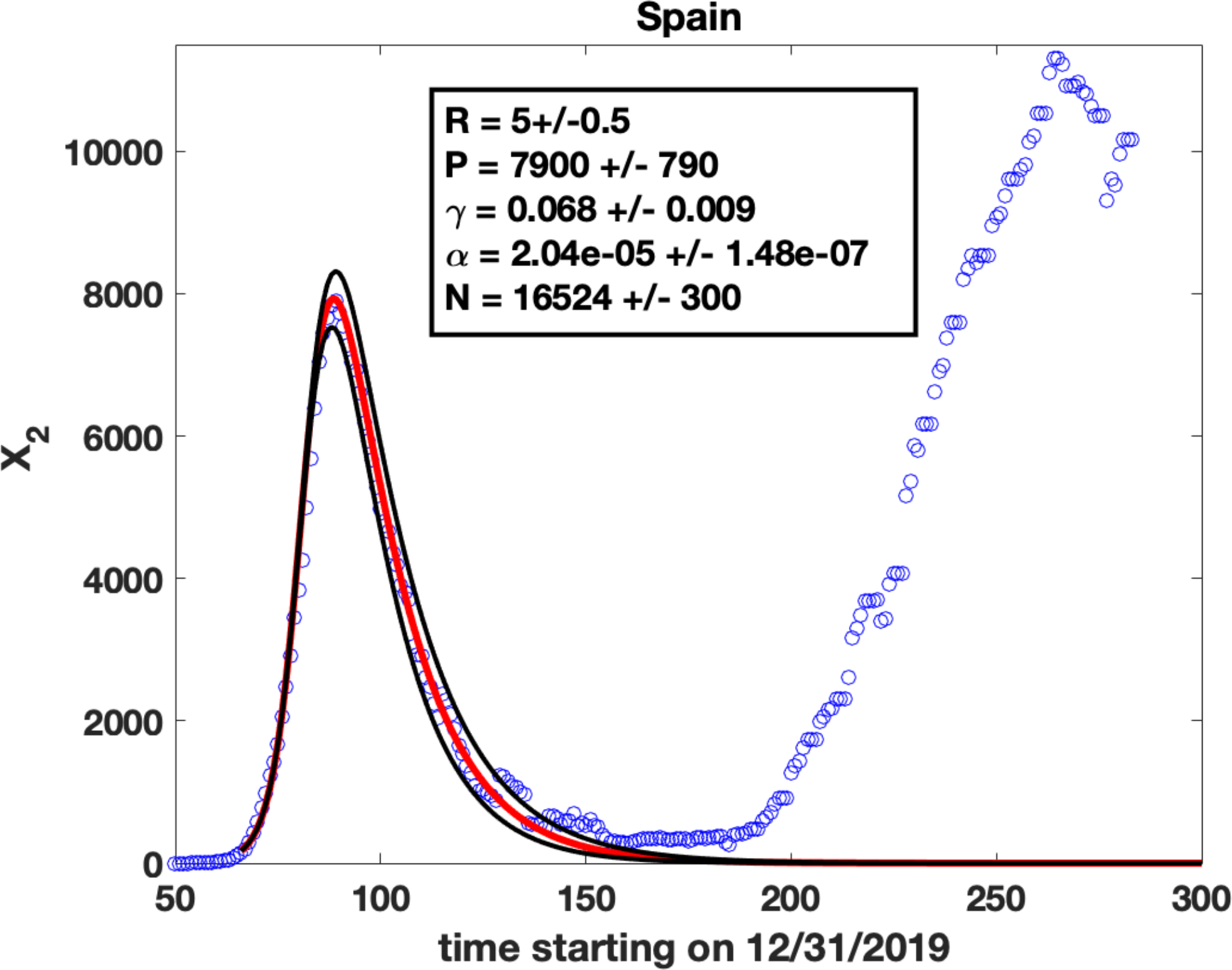
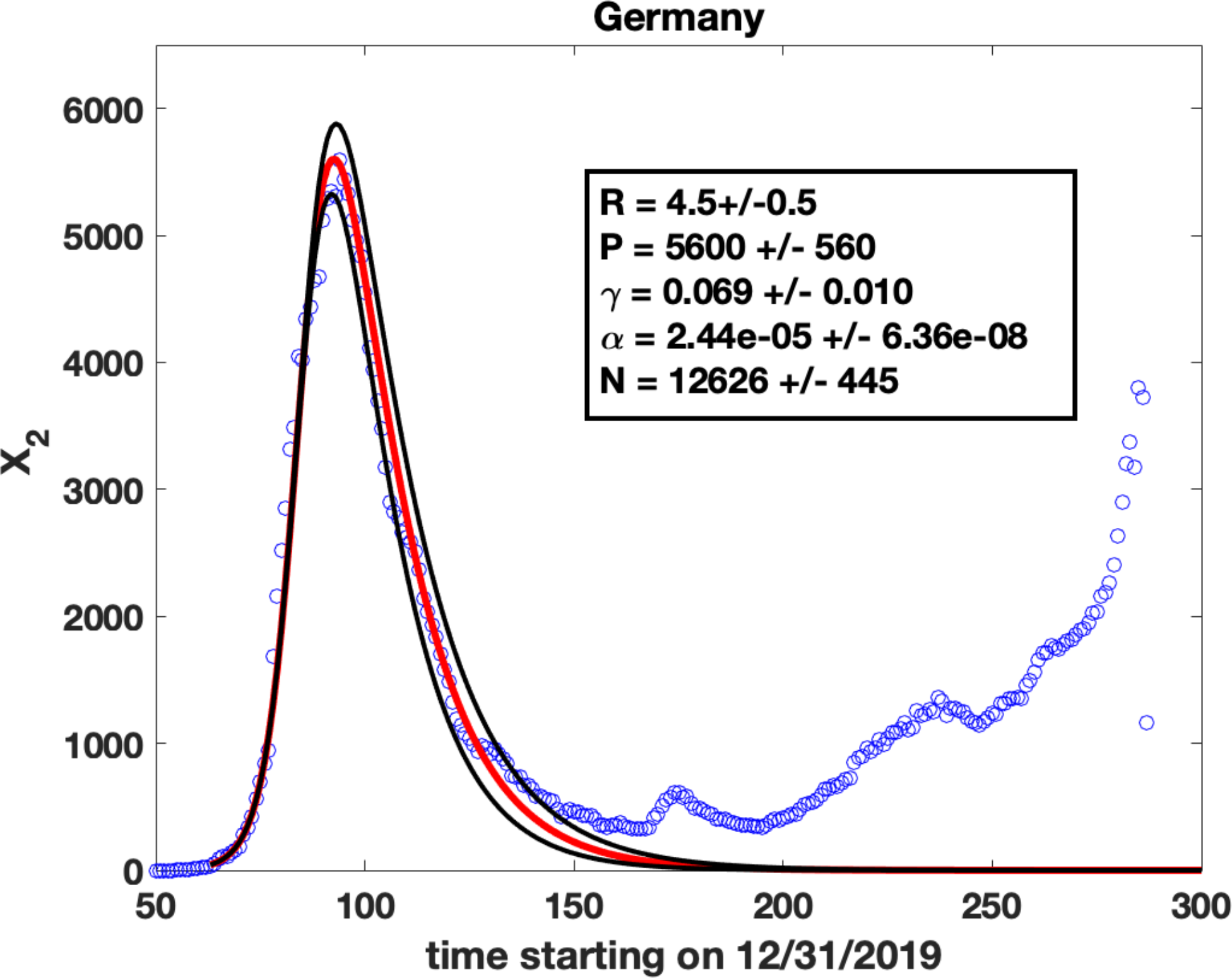
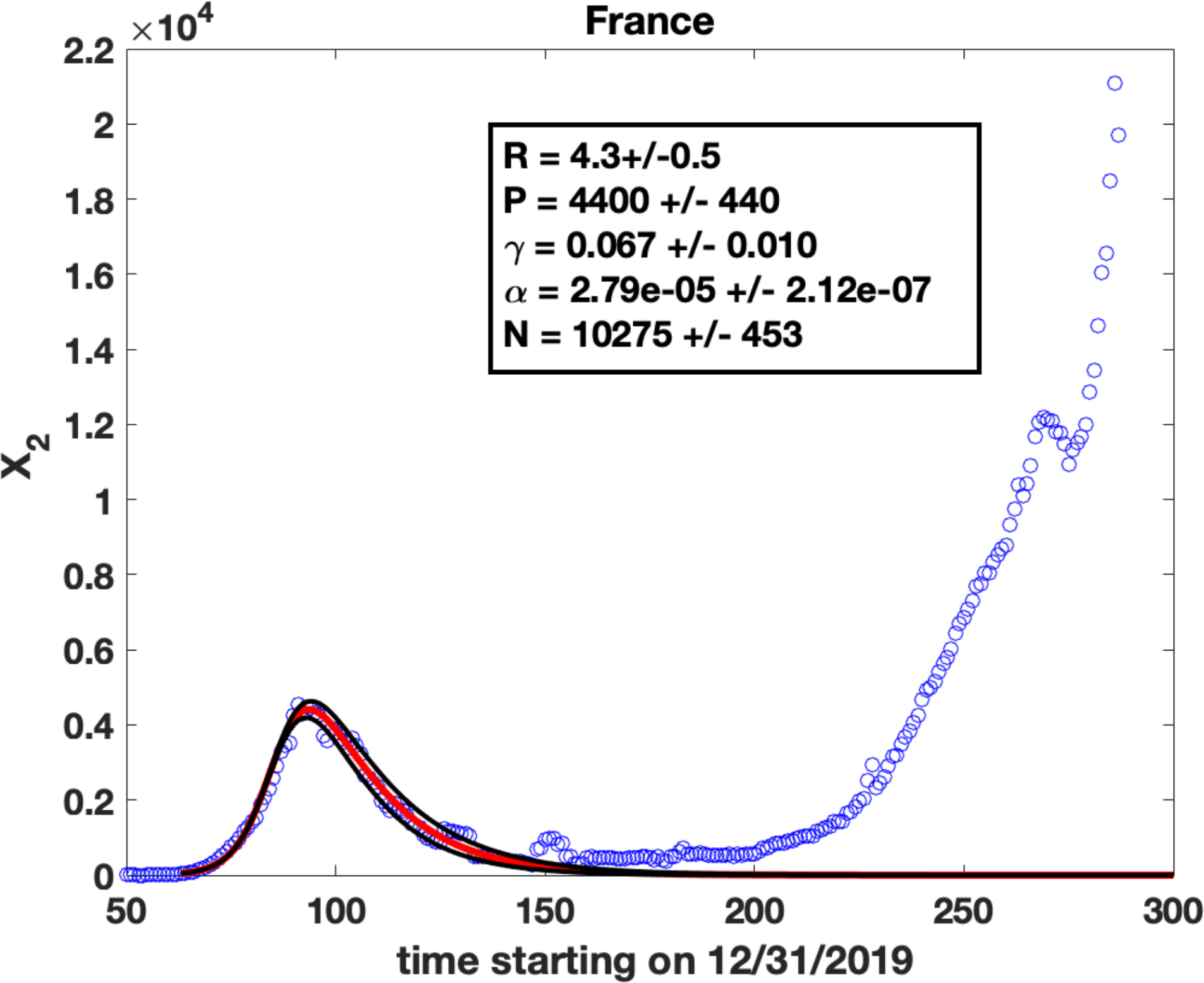
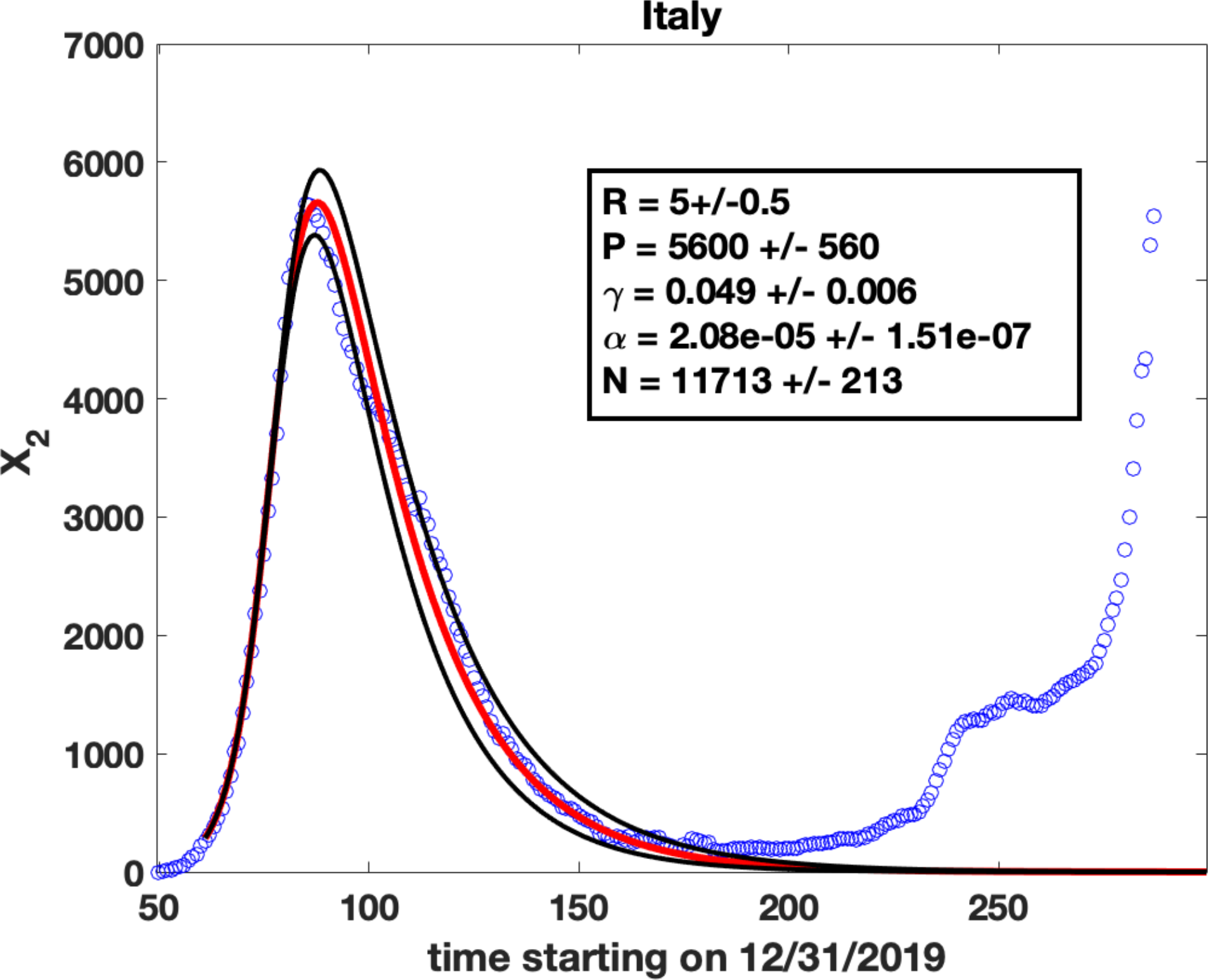
Parameter fits for the number of daily cases plotted on a linear scale: These plots are the same as those in Figure 2 except that the y axis is shown in a linear scale, rather than a log scale. This is to show the accuracy of the fits which are shown as solid lines. Blue circles are observed data for *X*_2_(*t*), the number of cases per day. Solid lines are fits obtained by solving (8) and (9) using the ODE solver *ode45* in Matlab. The values of the parameters are shown in the insets and represent the fit shown as the solid red line. The method used for the fits was to find ***γ***(*R* − 1) from the exponential rise in *X*_2_ at early times (Appendix A), estimate the peak value ***P*** of *X*_2_ (which gives the value of *N* using (15)) and find an *R* value that best fits the data (red solid line). The black lines represent solver results for R varying by 0.5 from the best fit value.

