## Appendix A for "Analysis of Covid-19 Data for Eight European Countries and the United Kingdom Using a Simplified SIR Model"

The basic equations are:

$$\frac{dX_1}{dt} = -\alpha X_1 X_2 \quad \text{(A1)}$$
$$\frac{dX_2}{dt} = \alpha X_1 X_2 - \gamma X_2 \quad \text{(A2)}$$

The boundary conditions at $t = 0$ are:
$X_1(0) = (N - a)$ and $X_2(0) = a$ (A3)

Given these boundary conditions, $X_1(t)$ and $X_2(t)$ are related by (see (13) in Methods):

$$X_2(t) = N - a + \frac{N}{R}\log\left(\frac{X_1(t)}{N-a}\right) - X_1(t) \quad \text{(A4)}$$

For small t, we can expand the log in (A4) as follows:

$$\log\left(\frac{X_1(t)}{N-a}\right) = \log\left[1 - \left(1 - \left(\frac{X_1(t)}{N-a}\right)\right)\right] \sim -\left(1 - \left(\frac{X_1(t)}{N-a}\right)\right) \quad \text{(A5)}$$

which, when substituted into (A4) gives, after some simple algebra,

$$X_2(t) = \frac{R-1}{R}\left[N - X_1(t)\left(1 - \frac{a}{(R-1)N}\right)\right] \sim \frac{R-1}{R}\left[N - X_1(t)\right] \quad \text{(A6)}$$

Substituting these into (A1), we get the Logistic Equation:

$$\frac{dX_1}{dt} = -\gamma(R-1)X_1(t)\left[1 - \frac{X_1(t)}{N}\right] \quad \text{(A7)}$$

whose solution, with the boundary condition (A3) is:

$$X_1(t) = \frac{N}{\left[1 + \frac{a}{N-a}e^{\gamma(R-1)t}\right]} \quad \text{(A8)}$$

For small t, we can expand the denominator to get:

$$X_1(t) \sim N\left[1 - \frac{a}{N-a}e^{\gamma(R-1)t}\right], \text{ t small} \quad \text{(A9)}$$

Substituting into (A6) gives,

$$X_2(t) \sim \frac{(R-1)aN}{R(N-a)}e^{\gamma(R-1)t} \quad \text{(A10)}$$

This analysis shows that for small t, $X_1(t)$ decreases exponentially from its initial value of $(N - a)$ and this decrease feeds an exponential increase in $X_2(t)$.

(A10) also shows that the coefficient of $t$ in the exponent of $X_2(t)$ is $\gamma(R-1)$. (A11)
